# Genetic evidence for causal relationships between age at natural menopause and the risk of aging-associated adverse health outcomes

**DOI:** 10.1101/2022.01.26.22269835

**Authors:** Joanna Lankester, Jin Li, Elias Levy Itshak Salfati, Marcia L. Stefanick, Kei Hang Katie Chan, Simin Liu, Carolyn J. Crandall, Shoa L. Clarke, Themistocles L. Assimes

**Affiliations:** Department of Medicine, Stanford University School of Medicine; VA Palo Alto Health Care System, Palo Alto, CA; Thermofisher Scientific; The Scripps Research Institute; Department of Obstetrics and Gynecology, Stanford University School of Medicine; Departments of Biomedical Sciences and Electrical Engineering in the City University of Hong Kong; Center for Global Cardiometabolic Health, Brown University; Department of Epidemiology, School of Public Health, Brown University; Department of Medicine, Alpert School of Medicine, Brown University; Department of Medicine, David Geffen School of Medicine at University of California, Los Angeles

## Abstract

**Background:** A later age at natural menopause (ANM) has been linked to several aging-associated traits including an increased risk of breast and endometrial cancer and a decreased risk of lung cancer, osteoporosis, and Alzheimer disease. However, ANM is also related to several proxies for overall health that may confound these associations.

**Methods:** We investigated the causal association of ANM with these clinical outcomes using Mendelian randomization (MR). Participants and outcomes analyzed were restricted to post-menopausal females. We conducted a one-sample MR analysis in both the Women’s Health Initiative (WHI) and the UK Biobank (UKB). We further analyzed and integrated several additional datasets of post-menopausal women using a two-sample MR design. We used up to 55 genetic variants previously discovered to be associated with ANM as our instrumental variable.

**Results:** A five year increase in ANM was causally associated with a decreased risk of osteoporosis (OR=0.80 [0.70, 0.92]) and fractures (OR=0.76 [0.62, 0.94]) as well as an increased risk of lung cancer (OR=1.35 [1.06, 1.71]). Other associations including atherosclerosis related outcomes were null.

**Conclusions:** Our study confirms that the decline in bone density with menopause causally translates to fracture and osteoporosis. Additionally, this is the first causal epidemiologic analysis to our knowledge to find an increased risk of lung cancer with ANM. This finding is consistent with molecular and epidemiologic studies suggesting estrogen dependent growth of lung tumors. Randomized controlled trials of anti-estrogen therapies in the prevention or treatment of lung cancer should be considered if additional MR studies are confirmatory.

**Key Messages:**

- As in prior literature, the age of natural menopause (ANM) was observationally associated with increased risk of breast cancer, endometrial cancer, and ovarian cancer, and with a decreased risk of lung cancer, coronary heart disease, ischemic stroke, fracture, osteoporosis, and Alzheimer disease in the Women’s Health Initiative and UK Biobank.
- However, these associations may be confounded by overall markers of health, such as smoking, so we used a genetic instrument variable to look at the causality of ANM on these adverse outcomes using Mendelian randomization.
- A five year increase in ANM was causally associated with decreased risk of fracture and osteoporosis, but with an increase lung cancer.
- This increase in ANM was not significantly associated with other outcomes; notably, there was no causal association of ANM with coronary heart disease or ischemic stroke.
- Given the increase in lung cancer risk and prior molecular studies linking lung cancer to estrogen receptor expression, randomized controlled trials of anti-estrogen therapies for prevention or treatment of lung cancer should be considered, should these results be replicated in additional studies.

## Introduction

The age of natural menopause (ANM) has been linked to several aging-associated traits, including oncologic, cardiovascular, musculoskeletal, and neurocognitive related adverse health outcomes. Among oncologic outcomes, later menopause has been consistently associated with an increased risk in breast^1, 2^ and endometrial cancers^3, 4^ as well as a decreased risk in lung cancer^5, 6^. Associations with ovarian cancer have been less consistent^7^. For aging-associated traits unrelated to cancer, an older ANM has been consistently associated with a reduced risk of coronary heart disease (CHD) and ischemic stroke^8-11^ as well as a higher bone mass and a lower risk of fracture and of osteoporosis defined by bone mineral density T-score^12, 13^. Associations with cognition and dementia have been less consistent^14^. A strong biologic basis exists to explain some of these associations including for breast cancer, endometrial cancer, and osteoporosis^15-18^. The biological basis for the remaining associations, including a protection against lung cancer, and dementia, remains circumstantial.

Residual confounding may be responsible for at least some of the associations between ANM and aging-associated outcomes. For example, a later ANM has been linked to a lower rate of smoking, higher education, higher income, and higher physical activity^8, 19^, all of which affect the risk of several chronic diseases. Mendelian randomization (MR) represents a well-established approach to guard against residual confounding^20^. A recent large scale GWAS of ANM in about 200,000 women leveraged up to 290 genetic variants as instruments to conduct a two-sample MR between ANM and multiple health traits^21^. The investigators detected causal associations between ANM and an increased risk of breast and endometrial cancer, as well as a lower risk of reduced bone mass and fractures^21^. Less robust causal associations were detected with ovarian cancer and Type 2 diabetes and no association was detected for CHD^21^. An important limitation of this two-sample MR study was the use of publicly available summary statistics for GWAS as convenience datasets. Several of these datasets incorporated a large fraction of men (e.g., CHD) and/or a substantial number of premenopausal women (e.g., breast, endometrial, ovarian cancers, multiple cardiometabolic risk factors, and fractures) making MR inference much less reliable. Another limitation was the lack of study of the lung cancer outcome^21^.

Here, we overcome these limitations by first conducting a one-sample MR study within the Women’s Health Initiative (WHI) and the UK Biobank (UKB) using individual-level data, restricting to postmenopausal women with documented non-surgical menopause, and including the lung cancer outcome. We augment this one-sample analysis with additional publicly available individual-level datasets using a 2-sample framework but only after filtering out men as well as women with outcomes of interest occurring before menopause.

## Methods

### Datasets

A detailed description of the WHI and UKB study designs have previously been published^22-24^. The WHI study population consisted of post-menopausal women who had enrolled in the study between 1993 and 1998. The UKB data consisted of women recruited between ages 37-73 who had enrolled between 2006 and 2010. Among the data collected, our study utilized data from questionnaires, anthropometric measurements, and outcome data collected from self-report, primary care, hospital records, and death records. Data from both sources included postmenopausal women of European ancestry who had undergone natural menopause (**eText Items 2, 6**). We included for replication any available datasets in populations of European ancestry with a reported age of menopause variable and the outcome of interest (**eTable Item 1, eText Item 6**) which were downloaded from the NIH database of Genotypes and Phenotypes (dbGaP) and from the European Genome-Phenome Archive (EGA).

### Definitions of exposure and outcomes

We defined the age of natural menopause (ANM) in WHI as the self-reported age one year past the last menstrual period (LMP) for those who underwent non-surgical menopause, i.e., excluding women with a history of bilateral oophorectomy. Women who had undergone hysterectomy in the absence of bilateral oophorectomy had an estimated ANM based on their responses to questions about other menopausal symptoms (**eText Item 2**). In UKB, we defined ANM from the baseline questionnaire data as the self-reported age that menopause occurred in women without history of bilateral oophorectomy. Other menopause-related variables were not included in the questionnaire, and ANM was therefore not available for women who underwent hysterectomy prior to menopause. We additionally excluded those with these surgeries prior to 2 years after the reported age of menopause to mitigate against recall errors. In other datasets downloaded via dbGaP, we used the provided ANM or derived age of ANM from the same surgical criteria, where available. If the menopause phenotype was unavailable in these other datasets, we limited analyses to events occurring past the age of 55 (cases) or to women over the age of 55 (controls).

Outcomes in WHI were extracted for adjudicated cancers (breast, endometrial, ovarian, and lung cancers), adjudicated fractures, incident coronary heart disease and ischemic stroke, and self-reported osteoporosis and Alzheimer disease (**eTable Item 3b**). In UKB, cancers were extracted from the UK cancer registry. Fracture, osteoporosis, and Alzheimer disease were extracted from the first occurrences data, which reports the first date each ICD code was found in hospital, primary care, or death records, or in self-reported data from the intake survey (with the majority of our outcomes from hospital records). CVD outcomes were extracted from a combination of first occurrences and raw hospital data (**eTable Item 3d**). We restricted analyses to incident cases.

### Covariates

Covariates in observational analysis (**eTable Items 3a & 3c**) were age at enrollment, body mass index (BMI), status of oophorectomy and hysterectomy, status of menopause hormone therapy (MHT), smoking status, alcohol consumption, energy expenditure from exercise (WHI only, in weekly kcal/kg), and socioeconomic status (for WHI, education and family income; for UKB, Townsend index). We additionally adjusted for parity for estrogen-related cancers; systolic blood pressure (BP), hypertension, and diabetes for CVD outcomes; history of fracture and vitamin D (WHI - dietary; UKB - serum measurement) for fracture and osteoporosis outcomes; and baseline diabetes and history of stroke or transient ischemic attack for Alzheimer disease.

### Instrumental variable

Our instrumental variables (IV) for MR analysis were up to 55 single nucleotide polymorphisms (SNPs) previously discovered through a GWAS of ∼70,000 women of European ancestry (**eTable Item 4-5**)^25^ independent of all datasets analyzed in this study. We included all SNPs with a consistent direction of effect on ANM that were directly measured or were imputed on >90% of samples within each dataset. We assessed the strength of these instruments by measuring the F statistic of the association between a weighted genetic score constructed from these 55 SNPs with ANM in the WHI and the UKB datasets, with weights from the discovery GWAS. We also checked for the association between potential confounders and the IV to test for the second assumption of MR. Where significant associations existed, we conducted sensitivity analyses using multivariable MR to ensure that the third assumption of MR was not violated with a pathway around the exposure via these confounders, thus adjusting for the IV-MHT relationship for all outcomes and additionally for the IV-blood pressure relationship for coronary heart disease, stroke, and Alzheimer disease outcomes. Importantly, we did not use the larger set of 290 SNPs and insertion/deletions reported in the more recent larger GWAS of about 200,000 women^21^ as an instrument, as over half of the women included in that study were UK Biobank participants. Selection of this set of SNPs could result in an overfitting of instruments and exacerbation of weak instrument bias^26^ in our study which uses the UK Biobank as a primary dataset for our one-sample MR.

### Statistical analyses

We calculated summary statistics for the exposure, outcomes, and covariates for all subjects included in observational analyses, as well as for the subset with measured genotypes who were included in MR analyses. Next, we generated observational associations between ANM and each outcome in both WHI and UKB using logistic regression and adjusting for general and outcome-specific covariates. Observational associations from both cohorts were then combined using a random-effects meta-analysis. Our primary MR analysis was a random effects inverse variance weighted (IVW) method. This method combines the causal ratio estimates from each variant according to the variance on those estimates, where the ratio is computed as the outcome-IV association estimate divided by the exposure-IV association estimate. We adjusted each association for age at enrollment. We then combined the IVW results for each outcome in WHI and the UKB through a random effects meta-analysis. We further conducted two-sample IVW analyses from external publicly available datasets for outcomes where such datasets were available and premenopausal women could be reliably excluded and further combined these results with our one-sample MR analyses through a random-effects meta-analysis. We used an adjusted false discovery rate test to adjust for multiple outcomes testing. We conducted MR sensitivity analyses using weighted median analysis and MR-Egger and meta-analyzed these results between datasets. We also conducted MR-PRESSO analyses for the two-sample MR analyses and meta-analyzed any resulting outlier-corrected results with the main analyses. Analyses were performed using Stata/SE 13.1 (StataCorp LP, College Station, TX), R (https://cran.r-project.org/), and plink 2.0 (https://www.cog-genomics.org/plink/2.0/).

## Results

Menopausal women eligible for the study comprised 106,853 for the WHI observational analysis, 19,543 for the WHI MR analysis, and 95,464 for both UKB analyses (**Table 1**). Mean self-reported ANM was one year lower for WHI (49.3) compared to UKB (50.3). The cohorts were similar in BMI, smoking rates, and prevalence of diabetes at enrollment. The UKB had substantially lower rates of baseline hysterectomy, largely due to exclusions based on fewer survey questions related to menopause, and substantially lower rates of MHT use, largely related to differences in study design and years of data collection.

**Table 1.** Characteristics of Women’s Health Initiative and UK Biobank participants included in observational and one-sample instrumental variable (IV) analysis.

|  |  |  |  |
| --- | --- | --- | --- |
|  | Women's Health Initiative |  | UK Biobank |
|  | Observational analysis<br>(n=106853) | IV analysis<br>(n=19543) | Observational and<br>IV analysis<br>(n=95464) |
| **Quantitative variables** | mean(SD) or median (1st, 3rd quartile) |  |  |
| *Main exposure* |  |  |  |
| ANM (years) | 49.1 (5.9) | 49.3 (5.9) | 50.3 (4.5) |
| *Other* |  |  |  |
| age (years) | 63.6 (7.2) | 65.7 (6.9) | 60.6 (5.4) |
| BMI (kg/m<sup>2</sup>) | 27.5 (5.7) | 27.9 (5.7) | 27 (5.0) |
| systolic BP (mmHg) | 128.6 (19.2) | 131 (19.5) | 140.5 (20.3) |
| Townsend index |  |  | -1.7 (2.8) |
| education | 7.4 (1.8) | 7.2 (1.8) |  |
| family income | 4.3 (1.8) | 4.1 (1.7) |  |
| alcohol (drinks/week) | 2.4 (4.9) | 2.7 (5.3) | 5.4 (6.5) |
| PA (MET hours/week) | 9.3 (2.5, 18.8) | 8.3 (2.3, 17.5) |  |
| dietary Vitamin D (mcg) | 3.8 (2.4, 5.7) | 3.8 (2.4, 5.7) |  |
| vitamin D (nmol/L) |  |  | 50.6 (20.6) |
| number of pregnancies / parity | 2.6 (1.7) | 2.7 (1.7) | 1.9 (1.1) |
| WHO fracture risk score | 9.3 (6.5, 14.3) | 10.6 (7.5, 15.7) |  |
| **Binary variables** | n (%) |  |  |
| HT trial participant | 8655 (8.1) | 4534 (23.2) |  |
| Ca<sup>+</sup> & Vit. D trial participant | 11754 (11.0) | 3733 (19.1) |  |
| hormone use ever | 61868 (57.9) | 9674 (49.5) |  |
| takes MHT |  |  | 4409 (4.6) |
| current smoking | 6945 (6.5) | 1446 (7.4) | 7480 (7.8) |
| bilateral oophorectomy /<br>oophorectomy hx | 6091 (5.7) | 919 (4.7) | 1723 (1.8) |
| hysterectomy | 31094 (29.1) | 5257 (26.9) | 2886 (3.0) |
| hypertension | 23615 (22.1) | 4788 (24.5) | 45596 (47.8) |
| diabetes / T2DM | 3312 (3.1) | 762 (3.9) | 3172 (3.3) |
| broken bone | 42955 (40.2) | 8091 (41.4) |  |
| osteoporosis | 8441 (7.9) | 1524 (7.8) |  |
| stroke or TIA | 2885 (2.7) | 625 (3.2) | 1350 (1.4) |
| BMI: body mass index, BP: blood pressure, PA: physical activity, Ca<sup>+</sup>: Calcium, Vit. D: Vitamin D, WHO: World Health Organization, HT: Hormone Therapy, MHT: Menopausal hormone therapy, T2DM: Type 2 diabetes |  |  |  |

Observational analyses in the WHI and UKB combined linked a 5-year increase in ANM to a higher risk of breast, endometrial, and ovarian cancers, and a lower rate of lung cancer, CHD, ischemic stroke, and Alzheimer disease (**Figure 1, eTable Item 7**). ANM was also inversely associated with the risk of fracture and osteoporosis with the former having borderline significance (**Figure 1, eTable Item 7**).

**Figure 1:**
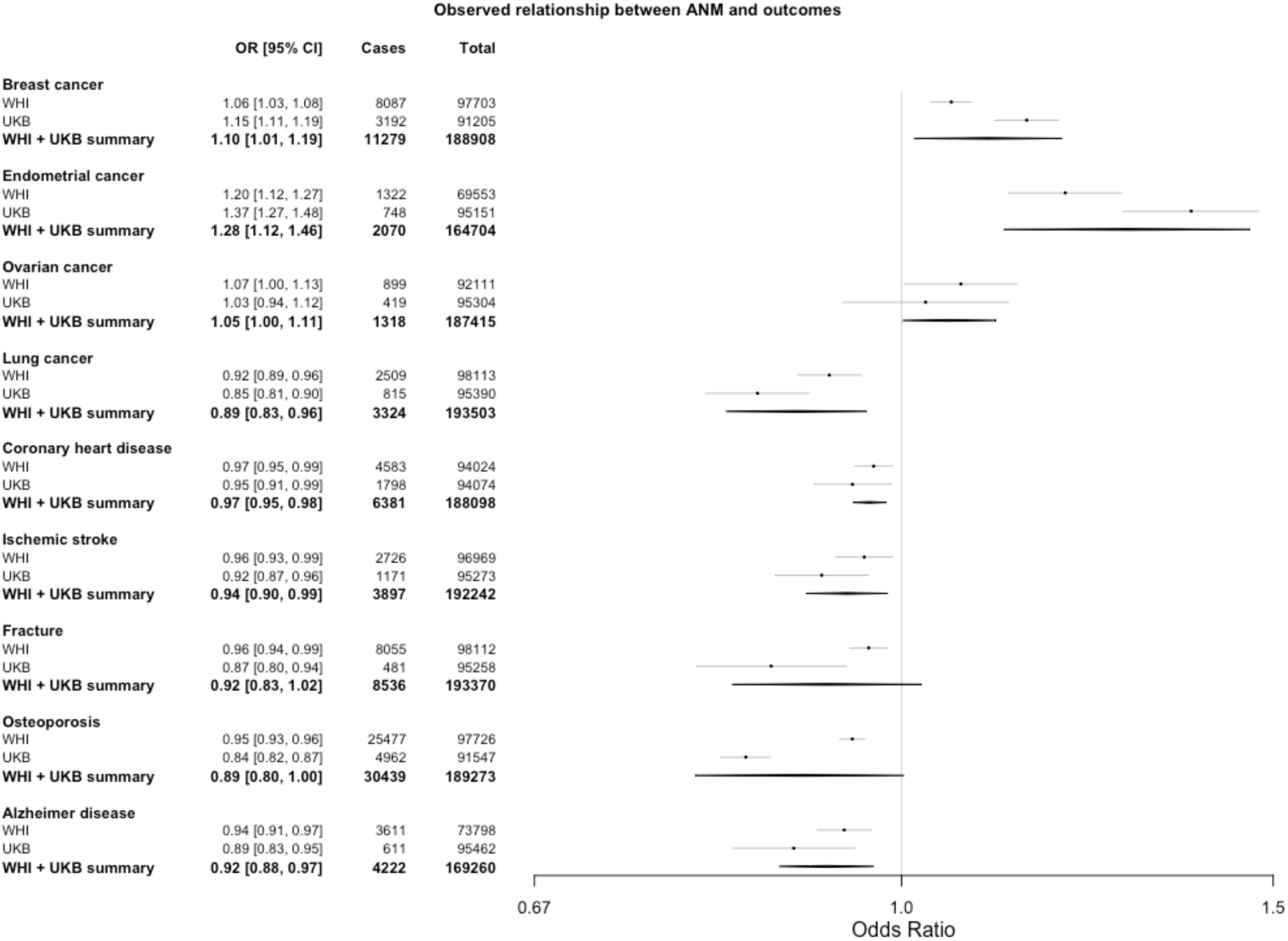
Results from the meta-analyzed observational logistic regression analyses of association between age of natural menopause and outcomes in WHI and UKB. Odds ratio for disease given a 5-year increase in age of natural menopause. Confidence interval, number of individuals who were cases for the disease in each dataset and analysis, and total number of individuals (cases + controls) from each dataset and analysis are also shown. ANM = age of natural menopause; OR = odds ratio; CI = confidence interval; WHI = Women’s Health Initiative; UKB = UK Biobank.

We found a weighted genetic risk score constructed with the ANM IV SNPs to be strongly associated with an older ANM in both WHI (F statistic=346, R^2^=1.8%) and UKB (F statistic=5062, R^2^=5.0%). The same score was not associated with any other baseline characteristics in WHI except for a nominal association with an indicator for participation in a WHI hormone trial (per 1 standard deviation increase in genetic risk score, OR=0.96, 95% confidence interval [0.93, 1.00]) and a history of a broken, fractured, or crushed bone (OR=0.97 [0.94, 1.00]). In UKB, we found the genetic risk score associated only with baseline characteristics of taking MHT (OR=0.89 [0.86,0.92]), age at enrollment (beta = 0.38 years [0.35, 0.42]), and systolic blood pressure (beta = 0.53 mmHg [0.39, 0.66]) (**eFigure Item 8**).

Our main MR results from the meta-analysis of WHI and UKB with external datasets showed ANM was causally associated with an increased risk of lung cancer (OR=1.35 [1.06,1.71] for each 5-year increase in ANM), in contrast to the protective effect in the observational analysis. The MR results also showed a causally protective effect for fracture (0.76 [0.62, 0.94]) and osteoporosis (0.80 [0.70, 0.92]) consistent with the protective effects in the observational analysis (**Figure 2, eTable Item 9**). The causal associations for lung cancer, osteoporosis, and fracture remained significant for multiple testing with a false discovery rate of 0.05 (**eTable Item 10**). Sensitivity analyses showed similar point estimates for weighted median and MR-Egger analyses with generally wider confidence intervals, retaining significance in weighted median analysis for osteoporosis only (OR=0.80 [0.68, 0.94]) **(eTable Item 9)**. ANM was not significantly associated with breast, endometrial, or ovarian cancers, CHD, ischemic stroke, or Alzheimer disease in the primary analysis. However, a strong trend toward significance for increase risk was observed for breast cancer in the overall sample as well as for endometrial cancer in our one sample MR meta-analysis restricted to WHI and UKB alone (**Figure 2, eTable Item 9**). Age at enrollment was already adjusted for in the MR analyses, and a multivariable MR adjusting for MHT and, where applicable, blood pressure gave similar results to the main MR results (**eFigure Item 11**). MR-PRESSO found 2 outlier SNPs in only one dataset (WTCCC2) which did not qualitatively influence the meta-analysis outcome (**eTable Item 12**).

**Figure 2:**
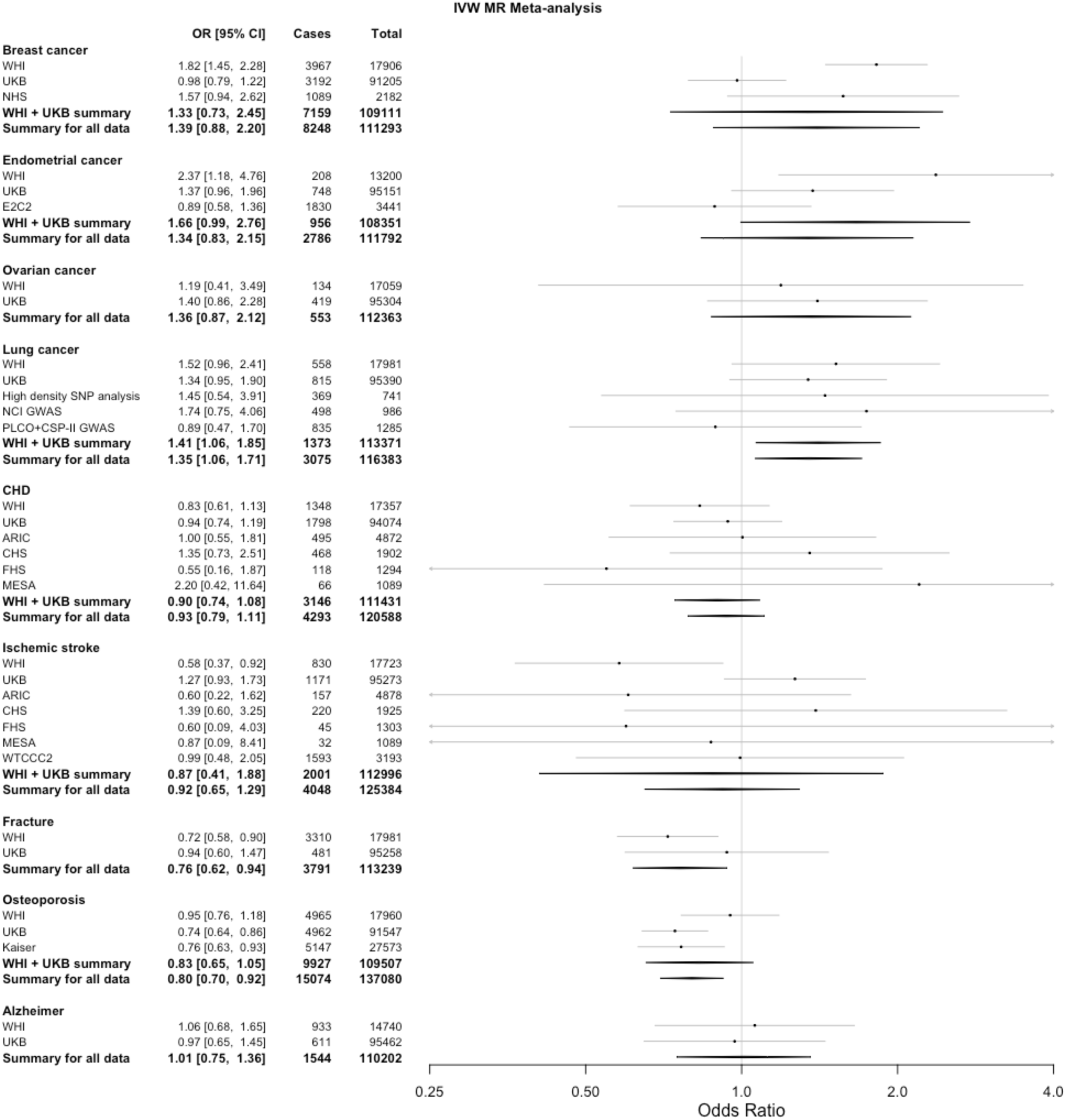
Results from the inverse variance weighted Mendelian randomization meta-analysis. Odds ratio for disease given a 5-year increase in age of natural menopause based on a causal genetic instrument variable analysis. Confidence interval, number of individuals who were cases for the disease in each dataset and analysis, and total number of individuals (cases + controls) from each dataset and analysis are also shown. IVW = inverse-variance weighted; MR = Mendelian randomization; OR = odds ratio; CI = confidence interval; WHI = Women’s Health Initiative; UKB = UK Biobank; NHS = Nurses Health Study; E2C2 = Epidemiology of Endometrial Cancer Consortium; High density SNP analysis = High Density SNP Association Analysis of Lung Cancer; NCI GWAS = National Cancer Institute (NCI) Genome Wide Association Study (GWAS) of Lung Cancer in Never Smokers; PLCO+CSP-II GWAS = The Prostate, Lung, Colon, Ovary Screening Trial + Cancer Prevention Study II); ARIC =The Atherosclerosis Risk in Communities Study; CHS = Cardiovascular Health Study; FHS = Framingham Heart Study; MESA = Multi-Ethnic Study of Atherosclerosis; WTCCC2 = Wellcome Trust Case Control Consortium 2.

## Discussion

We estimated causal associations between ANM and several aging-associated traits using the principal of MR in the WHI, the UK Biobank and several other datasets. Our principal findings are a causal decrease in the risk of fracture and osteoporosis with an increased age of menopause (ANM), a lack of causal association between ANM and atherosclerotic cardiovascular diseases (ASCVD) of CHD and ischemic stroke, and a causal increase in the risk of lung cancer. Of these three causal associations, only the findings related to bone health were consistent with findings from the analogous observational analysis. While a lack of a causal protective effect between ANM findings for CHD and stroke may be expected given results of recent randomized control studies (RCTs) of MHT, the causal increased risk for lung has never been previously reported. Our study is the first to our knowledge to study the causal relationship between ANM and many of these adverse outcomes using an MR analysis strictly limited to postmenopausal women. This segmentation is important to ensuring accurate inference in an MR study of adverse health outcomes related to ANM.

The protective effect of ANM for fracture and osteoporosis is expected given the compelling basic science and clinical evidence linking these health traits. Menopause is accompanied with a well-documented accelerated rate of bone loss whose mechanism is supported by extensive experimental evidence demonstrating the adverse effects of estrogen deficiency on the basic multicellular units responsible for bone remodeling^18^. Current recommendations for prevention of osteoporosis include screening for postmenopausal women, and the WHO fracture risk assessment model considers early menopause a risk factor for osteoporosis^27, 28^. Furthermore, multiple RCTs have shown the value of menopausal hormone therapy (MHT) in the prevention of osteoporosis^29^. While this benefit appears to be maximized when MHT is instituted immediately after menopause, it is also observed among women who started MHT at much later age. Nevertheless, MHT is approved only for prevention of osteoporosis; bisphosphonates are the current recommendation for management of osteoporosis in both older men and women given the balance between other beneficial and adverse effects of MHT^27, 28^.

The convincingly absent causal effect of ANM on ASCVD despite a strong inverse observational association suggests a substantial role of residual confounding and does not support the hypothesis that adverse changes to lipid profiles attributed specifically to the menopausal transition and independent of age are responsible for a protection from adverse health related outcomes with an older ANM^30-32^. MHT was originally studied for lipid profile improvement with two randomized crossover studies of conjugated estrogens in postmenopausal women with or without hyperlipidemia finding modest reductions in LDL of 15 and 24%, respectively^33, 34^. However, the WHI trials found that MHT slightly increased the risk of both CHD and stroke though neither was significant after multiple test correction^29^. These null findings may be due to a combination of both positive and negative effects of oral MHT on lipids, coagulation, inflammation, and endothelial function^35, 36^.

Our study is the first to our knowledge to implicate a causal association between an older ANM and a higher risk of postmenopausal lung cancer. Our MR analyses for this outcome were directionally opposite of the statistically significant protective effect of ANM documented in our observational analyses as well as that in other studies. This notable inconsistency suggests the presence of very substantial confounding which could be driven by factors such as smoking, diet, and exercise^8, 19^. Opposing directions of observational and MR studies are not common but have been documented in other settings including with the moderate use of alcohol^37^. Although our study is the first to link lung cancer and ANM through MR, another MR study of lung function similarly found early menopause associated with poorer lung function in observational analysis, but later menopause associated with poorer lung function in genetically instrumented analysis^38^. Similar to other cancers such as breast and endometrial cancers, our findings suggest that a greater length of exposure to estrogen with a later ANM may promote the transformation of lung cells or the growth and spread of existing subclinical primary lung tumors. In support of this hypothesis are numerous molecular studies of lung cancer which have found that estrogen receptors (ER) are present in lung tumours^39^, that both ER-alpha and ER-beta expression in cytoplasm are associated with poor lung cancer prognosis (with mixed evidence for nuclear ER-beta)^39, 40^, and that suppression of each of these receptors reduces lung cancer proliferation in vitro^41^. Multiple clinical outcome studies among cancer patients also provide persuasive evidence in support of this hypothesis. For example, the use of estrogen plus progestin was associated with a statistically significant hazard ratio of death from lung cancer in the WHI trials^42^. Furthermore, women who smoke are at greater risk than men who smoke, and an estrogen by smoking interaction has been hypothesized to explain this trend^39, 43^. Finally, observational analyses of population cancer datasets have found that anti-estrogen therapies improve lung cancer-specific survival among subjects with lung cancer in the presence or absence of prior history of breast cancer^44, 45^. If widely replicated, our findings suggest that anti-estrogen therapies could be repurposed to treat or prevent lung cancers in women at high risk assuming well powered RCTs are able to prove their benefits in this context.

A principal strength of our study was the MR design we implemented. First, we used two large studies with extensive, reliable, and broad ascertainment of health outcomes to a conduct a comprehensive MR study involving multiple outcomes relevant to the ANM. The one-sample MR design implemented in the WHI and UK Biobank allowed us to analyze only postmenopausal women. While ANM was not always available in the additional replication cohorts, access to individual level data allowed for two-samples analyses that were still restricted to postmenopausal women. Lastly, we found that the IV we used was largely independent of baseline characteristics that could confound our associations. We adjusted associations for baseline variables found to be nominal associated with our IV or found more significant associations to be inconsequential to the final results as documented through our multivariable MR sensitivity analyses.

The main limitation of our MR study was the potential for inadequate power for some of our outcomes despite the large sample sizes overall. Power in MR studies of binary outcomes is a function of sample size, variance explained of the exposure by the IV, the proportion of cases, the Type-1 error rate specified, and the true underlying causal odds ratio which is often not known^46^. Such a lack of power may be responsible for some of the statistically insignificant associations we observed in our study including for breast and endometrial cancers where substantial basic science, observational, and RCT evidence exists implicating increased risk through the effects of prolonged exposure to either endogenous or exogenous estrogens on the cellular transformation of epithelial cells^15-17, 29, 47^. For both breast and endometrial cancers, we are reassured by the fact that our MR results showed either a strong trend towards association overall (breast cancer) or in our main one-sample analysis (endometrial cancer), and that others have found a nominally significant or strongly positive association with larger sample sizes even if premenopausal cancers may have been included in the analysis^21, 47, 48^. Our null association of ANM with Alzheimer disease has recently also been documented using MR in an independent study^49^. The healthy cohort effect in UKB is also a known weakness that may have limited the number of cases and the generalizability of findings^50^. A weakness in phenotype definition was that osteoporosis and Alzheimer disease were self-reported in WHI. However, the meta-analysis result still remained positive for osteoporosis, and the result for Alzheimer disease was nearly identical to that of UKB where cases were derived from hospital records. These self-reported phenotypes therefore didn’t change the overall findings. Lastly, this study included only participants of European ancestry not only because White women were the majority of participants in most of the cohorts we examined, but also because the instruments were discovered through GWAS in predominantly White women. The causal effects of ANM should also be studied in more diverse populations as the diversity of biobank studies increases.

In conclusion, we report for the first time that a later ANM may causally increase the risk of lung cancer despite decreasing the risk of osteoporosis and fracture. Our findings point to the need for further randomized controlled trials of anti-estrogen therapies for the prevention or the treatment of lung cancer among postmenopausal women should our results be replicated in additional population genetic datasets.

### Ethics statement

The WHI project was reviewed and approved by the Fred Hutchinson Cancer Research Center (Fred Hutch) IRB in accordance with the U.S. Department of Health and Human Services regulations at 45 CFR 46 (approval number: IR# 3467-EXT). Participants provided written informed consent to participate. Additional consent to review medical records was obtained through signed written consent. Fred Hutch has an approved FWA on file with the Office for Human Research Protections (OHRP) under assurance number 0001920.

The UK Biobank data was accessed under Application Number 13721. The Research Ethics Committee reference for UK Biobank is 16/NW/0274. The Stanford IRB reviewed the protocol and determined the research did not include human subjects as defined in 45 CFR 46, nor 21 CFR 56.

Use of dbGaP and EGA datasets was approved by Stanford University IRB protocol #40313. The dbGaP data was accessed under project #2638. The EGA data was accessed under request ID #3367.

## Data Availability

All results produced in the present study are contained in the manuscript.
All data sets used in this study are publicly available to researchers by creating an account for the relevant data sources and following posted instructions for obtaining access.
Primary data from Women's Health Initiative used for this analysis are available through dbGap and the WHI website:
https://www.ncbi.nlm.nih.gov/projects/gap/cgi-bin/study.cgi?study_id=phs000746.v3.p3
https://www.whi.org
Phenotypic WHI data alone are available through BioLINCC:
https://biolincc.nhlbi.nih.gov/home/
Data from the UK Biobank can be accessed through the website:
https://www.ukbiobank.ac.uk/
Data from other sources are available through the NCBI's database of Genotypes and Phenotypes (dbGaP) and the European Genomic Archive (EGA):
https://www.ncbi.nlm.nih.gov/projects/gap/cgi-bin/study.cgi?study_id=phs000280.v7.p1
https://www.ncbi.nlm.nih.gov/projects/gap/cgi-bin/study.cgi?study_id=phs000287.v7.p1
https://www.ncbi.nlm.nih.gov/projects/gap/cgi-bin/study.cgi?study_id=phs000893.v1.p1
https://www.ncbi.nlm.nih.gov/projects/gap/cgi-bin/study.cgi?study_id=phs000336.v1.p1
https://www.ncbi.nlm.nih.gov/projects/gap/cgi-bin/study.cgi?study_id=phs000007.v32.p13
https://www.ncbi.nlm.nih.gov/projects/gap/cgi-bin/study.cgi?study_id=phs000753.v1.p1
https://www.ncbi.nlm.nih.gov/projects/gap/cgi-bin/study.cgi?study_id=phs000788.v2.p3
https://www.ncbi.nlm.nih.gov/projects/gap/cgi-bin/study.cgi?study_id=phs000674.v3.p3
https://www.ncbi.nlm.nih.gov/projects/gap/cgi-bin/study.cgi?study_id=phs000209.v13.p3
https://www.ncbi.nlm.nih.gov/projects/gap/cgi-bin/study.cgi?study_id=phs000634.v1.p1
https://www.ncbi.nlm.nih.gov/projects/gap/cgi-bin/study.cgi?study_id=phs000147.v3.p1
https://ega-archive.org/datasets/EGAD00010000264

https://www.whi.org

https://biolincc.nhlbi.nih.gov/home/

https://www.ukbiobank.ac.uk/

https://www.ncbi.nlm.nih.gov/projects/gap/cgi-bin/study.cgi?study_id=phs000280.v7.p1

https://www.ncbi.nlm.nih.gov/projects/gap/cgi-bin/study.cgi?study_id=phs000287.v7.p1

https://www.ncbi.nlm.nih.gov/projects/gap/cgi-bin/study.cgi?study_id=phs000893.v1.p1

https://www.ncbi.nlm.nih.gov/projects/gap/cgi-bin/study.cgi?study_id=phs000336.v1.p1

https://www.ncbi.nlm.nih.gov/projects/gap/cgi-bin/study.cgi?study_id=phs000007.v32.p13

https://www.ncbi.nlm.nih.gov/projects/gap/cgi-bin/study.cgi?study_id=phs000753.v1.p1

## Acknowledgement

The WHI programs is funded by the National Heart, Lung, and Blood Institute, National Institutes of Health, U.S. Department of Health and Human Services through 75N92021D00001, 75N92021D00002, 75N92021D00003, 75N92021D00004, 75N92021D00005. We acknowledge the following WHI investigators—program office: Jacques Rossouw, Shari Ludlam, Joan McGowan, Leslie Ford, and Nancy Geller (National Heart, Lung, and Blood Institute, Bethesda, MD), clinical coordinating center: Garnet Anderson, Ross Prentice, Andrea LaCroix, and Charles L Kooperberg (Fred Hutchinson Cancer Research Center, Seattle, WA), investigators and academic centers: JoAnn E. Manson (Brigham and Women’s Hospital, Harvard Medical School, Boston, MA), Barbara V. Howard (MedStar Health Research Institute/Howard University, Washington, DC), Marcia L. Stefanick (Stanford Prevention Research Center, Stanford, CA), Rebecca Jackson (The Ohio State University, Columbus, OH), Cynthia A. Thomson (University of Arizona, Tucson/Phoenix, AZ), Jean Wactawski-Wende (University at Buffalo, Buffalo, NY), Marian Limacher (University of Florida, Gainesville/Jacksonville, FL), Jennifer Robinson (University of Iowa, Iowa City/Davenport, IA), Lewis Kuller (University of Pittsburgh, Pittsburgh, PA), Sally Shumaker (Wake Forest University School of Medicine, Winston-Salem, NC), Robert Brunner (University of Nevada, Reno, NV), Karen L. Margolis (University of Minnesota, Minneapolis, MN); WHI memory study: Mark Espeland (Wake Forest University School of Medicine, Winston-Salem, NC). Joanna Lankester was supported by the National Heart, Lung, and Blood Institute through grant number 5F32HL149254-02 and by a Big Data Scientific Training Enhancement Program (BD-STEP) fellowship through the Palo Alto Veteran’s Affairs.

## Items for supplement

1. Table: external datasets included
2. Text: Description of derived ANM phenotype
3. Tables: Definitions of covariates and outcomes in WHI and UKB datasets
4. Table: SNPs included in IV and summary statistics for relationship with ANM
5. Table: R-squared for imputation of SNPs in each data set.
6. Text: Description of genetic ancestry in various data sets
7. Table: Observational results
8. Figures: Associations of ANM and confounders with instrument variables
9. Table: MR results for main analysis and sensitivity analyses
10. Tables: Estimates, standard errors, and p values from IVW MR analysis, and false discovery rate results for multiple testing correction
11. Figure: Multivariate results vs. univariate results from MR in UK Biobank
12. Tables: MR-PRESSO analysis result and meta-analysis of MR-PRESSO with other data sets for ischemic stroke

