## Supplemental Information for "Genetic evidence for causal relationships between age at natural menopause and the risk of aging-associated adverse health outcomes"

[3]Thermofisher Scientific

[4]The Scripps Research Institute

[5]Department of Obstetrics and Gynecology, Stanford University School of Medicine

[6]Departments of Biomedical Sciences and Electrical Engineering in the City University of Hong Kong

[7]Center for Global Cardiometabolic Health, Brown University

[8]Department of Epidemiology, School of Public Health, Brown University

[9]Department of Medicine, Alpert School of Medicine, Brown University

[10]Department of Medicine, David Geffen School of Medicine at University of California, Los Angeles

### Supplemental Methods

#### Item 1.

Table. External datasets acquired from NIH database of Genotypes and Phenotypes (dbGaP) and from the European Genome-Phenome Archive (EGA) for two-sample mendelian randomization after restricting datasets to post-menopausal women. *Datasets without age of menopause available were filtered on age of diagnosis >55 years. NHS: This study included only postmenopausal cases in the initial stage (CGEMS - Cancer Genetic Markers of Susceptibility) and controls from NHS.

| Abbreviation in figure | Full name | Access number | Phenotype | Age of menopause available?* | Reference (PMID) |
|---|---|---|---|---|---|
| ARIC | The Atherosclerosis Risk in Communities (ARIC) Study | phs000280 | coronary heart disease, stroke | yes | 2646917 |
| CHS | Cardiovascular Health Study | phs000287 | coronary heart disease, stroke | yes | 1669507 |
| E2C2 | Epidemiology of Endometrial Cancer Consortium | phs000893 | endometrial | no | 19132539, 24096698 |
| FHS | Framingham Heart Study | phs000007 | coronary heart disease, stroke | yes | 14819398, 1208363, 17372189 |
| High density SNP analysis | High Density SNP Association Analysis of Lung Cancer | phs000753 | lung cancer | no | 18385676 |
| Kaiser | Kaiser Permanente Research Program on Genes, Environment, and Health (RPGEH) | phs000788 (phs000674) | osteoporosis | no | 21565264, 21903159, 26092718, 26092716 |
| MESA | Multi-Ethnic Study of Atherosclerosis | phs000209 | coronary heart disease, stroke | yes | 12397006 |
| NCI GWAS | National Cancer Institute (NCI) Genome Wide Association Study (GWAS) of Lung Cancer in Never | phs000634 | lung cancer | no | 31009812 |
| NHS | Nurses Health Study 2 | phs000147 | breast cancer | no | 17529973, 22037553 |
| PLCO + CSP-II GWAS | A Genome-Wide Association Study of Lung Cancer Risk | phs000336 | lung cancer | no | 19836008 |
| WTCCC2 | Wellcome Trust Case Control Consortium 2 | EGAD00010000264 | stroke | no | 24436234 |

#### Item 2.

**Definition of ANM**

Women's Health Initiative (WHI): Age of natural menopause was derived by WHI from questions related to menopause status and provided via the MENO variable which we used as the ANM in our analyses. Questions related to menopause status included "How old were you when you last had any menstrual bleeding?", "When was the last time you had any menstrual bleeding or spotting?", "How old were you when you first had symptoms such as hot flashes or night sweats?", "Did you ever have a hysterectomy?", "How old were you when you had your hysterectomy", "Did you ever have an operation to have one or both of your ovaries taken out?", and "How old were you when you had your last operation to remove an ovary?" For women with history of bilateral oophorectomy before menopausal symptoms began, their ANM is not available. For women with history of hysterectomy in the absence of bilateral oophorectomy before menopausal symptoms began, their ANM was estimated from the age of their menopausal symptoms instead of last menstrual bleeding. For the remaining women, their ANM was derived from the age of last menstrual bleeding and defined as the age of the participant after 1 year of amenorrhea.

UK Biobank (UKB): Women who indicated they had undergone menopause were asked: "How old were you when your periods stopped?" Their response was recorded in field 3581 labeled "age at menopause". Given the ambiguity of the question (i.e., they were not asked for age at last menstrual period), we took this field at face value. This field was collected from women of a variety of ages in the UK Biobank and therefore could have been many years post-menopause. To mitigate against recall error, we used a two-year buffer for definition of both surgical causes of period cessation and clinical outcome cases. Thus, women who had undergone bilateral oophorectomy (field 3882) at or before the age of field 3581 + 2 years were excluded. UKB did not collect data on other menopausal symptoms, so women who had a hysterectomy prior to menopause did not have an available age of menopause. Those with an age of hysterectomy (field 2824) at or before the age of field 3581 + 2 years were also excluded.

### Item 3.

Phenotypic definitions: unique identifiers for each variable used from Women's Health Initiative and UK Biobank. Retrievable from (https://sp.whi.org/researchers/data/SitePages/SAS%20Variables.aspx) and (https://biobank.ctsu.ox.ac.uk/crystal/search.cgi).

#### Item 3a.

Table. Definition of covariates in Women's Health Initiative data. (Documented variable order may differ from question number in form, given that some questions have subsections.) CT = Clinical trial; OS = Observational study.

| Covariate | Variable ID | Units | Form | Variable Order | Description |
|---|---|---|---|---|---|
| age at menopause | MENO | years | 31 | 63 | Age at which participant went through menopause. Computed using Form 31, questions 5, 6.1, 10, 10.1; Form 2, questions 7, 18, 18.2; Form 43 (age first used hormone replacement therapy). |
| age at enrollment | AGE | years | 2 | 6 | Derived from date of birth and date of enrollment |
| body mass index | BMI | kg/m^2 | 80 | 20 | Derived from measured height and weight at enrollment |
| systolic blood pressure | SYST | mmHg | 80 | 7 | Measured at baseline (question 6.1 and 6.2), averaged from two readings |
| education | EDUC | numeric category | 20 | 3 | ordinal variable with 11 categories (question 6) |
| family income | INCOME | numeric category | 20 | 25 | ordinal variable with 9 categories (question 11) |
| alcohol | F60ALCWK | number of servings per week | 60 | 32 | self-reported from food frequency questionnaire |
| physical activity | TEXPWK | MET-hours/week | 143 | 210 | Computed from form 143, questions 6, 6.1, 6.2, 7.1-7.6 |
| dietary vitamin D | F60VITD | mcg/day | 60 | 56 | self-reported from food frequency questionnaire |
| number of pregnancies | PARITY | number of pregnancies | 31 | 55 | Number of pregnancies lasting at least 6 months, computed from form 31, questions 7, 7.6, 7.7 |
| World Health Organization fracture risk | WHOFRAC | risk score | n/a | Risk scores 2 | Derived using the FRAX tool |
| current smoking | SMOKING | True/False | 34 | 48 | Computed from Form 34, questions 1, 1.2, and 1.5. Selected current smoking (True/False). |
| participated in menopausal hormone therapy (MHT) trial | HRTFLAG | True/False | Demographics and Study Membership (CT+OS) | 4 | n/a |
| hormone use ever | TOTH | True/False | 43 | 32 | hormone replacement therapy use ever, collected at baseline |
| participated in calcium and vitamin D trial | CADFLAG | True/False | Demographics and Study Membership (CT+OS) | 4 | n/a |
| bilateral oophorectomy | BOOPH | True/False | 31 | 60 | Computed from for 31, question 10. |
| hysterectomy | HYST | True/False | 2 | 22 | From form 2, question 18. |
| hypertension history | HTNTRT | True/False | 30 | 109 | Computed from Form 30, questions 7, 7.2, and 7.3. Three category variable on history of hypertension including information on current treatment. The three groups are never, currently untreated and currently treated hypertensive. |
| diabetes history | DIABTRT | True/False | 2 | 69 | Computed from Form 2, question 23, 23.4 and 23.6. Indicator for whether the participant has ever been treated for diabetes with pills or shots. |
| broken bone history | BKBONE | True/False | 30 | 92 | Self-reported at baseline from question 15 |
| osteoporosis history | OSTEOPOR | True/False | 30 | 16 | Self-reported at baseline from question 2 |
| stroke or TIA history | STROKE, TIA | True/False | 2 | 43, 45 | Self-reported as having been diagnosed by a doctor from questions 26, 27. |

Item 3b.

| Table. Definition of clinical outcomes in Women's Health Initiative (WHI) data. | | | | |
|---|---|---|---|---|
| **Outcome** | **Variable ID** | **Form** | **Variable Order** | **Description** |
| Breast cancer | BREAST | * | 97 | Situ Breast Cancer. All Deaths due to breast cancer are considered Invasive Breast Cancers. Breast Cancer is adjudicated for CT and OS participants. |
| Endometrial cancer | ENDMTRL | * | 124 | Defined as the first occurrence of Endometrial Cancer or a Death due to Endometrial Cancer. Endometrial Cancer is adjudicated for CT and OS participants. |
| Ovarian cancer | OVARY | * | 205 | Defined as the first occurrence of Ovarian Cancer or a Death due to Ovarian Cancer. Ovarian Cancer is adjudicated for CT and OS participants. |
| Lung cancer | LUNG | * | 160 | Defined as the first occurrence of Lung Cancer or a Death due to Lung Cancer. Lung Cancer is adjudicated for CT and OS participants. |
| Coronary heart disease | CHD | * | 16 | Defined as the first occurrence of Clinical MI, Definite Silent MI or a Death due to Definite CHD or Possible CHD. Clinical MI and Death were adjudicated for CT and OS ppts through Ext1. In Ext2 they are only adjudicated for the Medical Record Cohort. |
| Ischemic stroke | STRKISCH | * | 40 | Defined as a Stroke where the diagnosis question indicates an iscemic stroke. Ischemic Stroke was adjudicated for CT and OS participants through Ext1. In Ext2 it is only adjudicated for the Medical Record Cohort. |
| Fracture | ANYFX | * | 280 | Defined as the first fracture of Hip, upper leg, pelvis, knee, lower leg, ankle, foot(not toe), tailbone, spine or back, lower arm or wrist, hand (not finger), elbow, upper arm or shoulder and other. |
| Osteoporosis | F33OSTEOP | ** | 43 | Not collected after WHI study closeout. |
| Alzheimer disease | F33ALZHEIMERS | ** | 71 | Combined from all three instances (CT/OS extension 1, CaD extension 1, and extension 1) |
| CT = Clinical Trial; OS = Observational Study; MI = myocardial infarction; CHD = coronary heart disease; Ext = extension study; CaD = Calcium and Vitamin D study; * = from Clinical Trial (CT) and Observational Study (OS) adjudicated outcomes; ** = self-reported outcomes for CT/OS through WHI Extension study 1. | | | | |

Item 3c.

| Table. Definition of covariates in the UK Biobank data. | | | |
|---|---|---|---|
| **Covariate** | **Variable ID** | **Units** | **Description** |
| age at menopause | 3581 | years | Details given in text and supplement text |
| age at enrollment | 21022 | years | Value provided as an integer |
| body mass index | 21001 | kg/m^2 | Derived from measured height and weight at enrollment |
| systolic blood pressure | 4080 | mmHg | Measured at enrollment |
| Townsend index (SES) | 189 | n/a | Numeric representation of material deprivation |
| weekly alcohol | 1558, 4462, 5364, 4429, 4418, 4451, 4407, 4440, 1588, 1578, 1608, 1568, 1598 | glasses of wine equivalent per week | Annual alcohol (g) was factored according to the list of all alcohol types typically consumed. This number was divided by 52 weeks and then converted into an equivalent number of glasses of wine. |
| serum vitamin D | 30890 | nmol/L | Laboratory value from serum drawn at enrollment |
| parity | 2734 | children | Number of live births |
| current smoking | 20116 | True/False | Self-reported |
| takes menopausal hormone therapy (MHT) | 6153 | True/False | Self-reported |
| bilateral oophorectomy | 2834 (status), 3882 (age, if applicable) | True/False | Details given in text and supplement text |
| hysterectomy | 3591 (status), 2824 (age, if applicable) | True/False | Details given in text and supplement text |
| hypertension history | 131286, 53 | True/False | True if date of 131286 (ICD I10 diagnosis) <= date of enrollment in field 53 |
| type II diabetes history | 2443, 4041, 20002, 20003, 2976, 2986, 30750 | True/False | Candidates included participants who had self-reported diabetes diagnosed by doctor that was not gestational only, took diabetes medications, or had an HbA1C that exceeded 48 mmol/mol. Those who had ever received a type 1 diabetes diagnosis code, who were diagnosed before age 35, or who required insulin within one year were then excluded (as clearly type 1 diabetes or potentially miscoded type 1 diabetes). |
| stroke or TIA history | 20002, 131366, 53 | True/False | Self-reported condition code 1081, 1082, or 1583; or, date of 131366 (ICD I63 diagnosis) preceeded date of enrollment in field 53 |

Item 3d.

| Table. Definition of outcomes in UK Biobank data. | | | | | |
|---|---|---|---|---|---|
| **Outcome** | **ICD-10 codes** | **ICD-9 codes** | **Procedure codes** | **Variable ID number (or variable column name, for hospital data)** | **Description** |
| Breast cancer | C50 | 174 | n/a | 40006, 40013 | Cancer registry |
| Endometrial cancer | C54.1 | 182 | n/a | 40006, 40013 | Cancer registry |
| Ovarian cancer | C56 | 183 | n/a | 40006, 40013 | Cancer registry |
| Lung cancer | C34 | 162 | n/a | 40006, 40013 | Cancer registry |
| Coronary heart disease | I20.0*, I21, I22 | 410, 411 | K49, K50, K75, K40-K46 | 131298, 131300, diag_icd10*,diag_icd9*, oper4* | First occurrences, Hospital raw data* |
| Ischemic stroke | I63 | 433, 434 | n/a | 131366, diag_icd10*,diag_icd9*, oper4* | First occurrences, Hospital raw data* |
| Fracture | M80 | n/a | n/a | 131962 | First occurrences |
| Osteoporosis | M80, M81 | n/a | n/a | 131962, 131964 | First occurrences |
| Alzheimer disease | F00, G30 | n/a | n/a | 130836, 131036 | First occurrences |

#### Item 4

Table. Single nucleotide polymorphisms reported by Day et al. in Pubmed ID 26414677 used as instrumental variables in Mendelian Randomization in this study.

| Locus (Closest gene) | SNP | Chr | Position (GRCh37) | Weight | Non effect Allele | Effect Allele (EA) | EA freq in WHI | β (SE) in WHI | F statistic in WHI | EA freq in UKB | β (SE) in UKB | F statistic in UKB |
|---|---|---|---|---|---|---|---|---|---|---|---|---|
| *RHBDL2, MYCBP* | rs4246511 | 1 | 39380385 | 0.22 | C | T | 0.31 | 0.14 (0.07) | 4.32 | 0.3 | 0.22 (0.02) | 97.97 |
| *RAD54L* | rs12142240 | 1 | 46747301 | 0.13 | T | C | 0.3 | 0.04 (0.07) | 0.46 | 0.31 | 0.15 (0.02) | 47.41 |
| *STX6* | rs1411478 | 1 | 180962282 | 0.13 | A | G | 0.59 | 0.21 (0.06) | 11.68 | 0.6 | 0.14 (0.02) | 42.27 |
| *EXO1* | rs2236918 | 1 | 242017826 | 0.15 | C | G | 0.55 | 0.01 (0.06) | 0.03 | 0.55 | 0.13 (0.02) | 39.13 |
| *BRE, GTF3C2,EIFB4* | rs704795 | 2 | 27716494 | 0.16 | A | G | 0.62 | 0.22 (0.06) | 12.35 | 0.6 | 0.19 (0.02) | 82.94 |
| *MSH6* | rs1800932 | 2 | 48018081 | 0.17 | A | G | 0.18 | 0.18 (0.08) | 5.31 | 0.19 | 0.21 (0.03) | 63.8 |
| *TLK1* | rs930036 | 2 | 171941018 | 0.19 | A | G | 0.62 | 0.11 (0.06) | 3.05 | 0.62 | 0.17 (0.02) | 66.84 |
| *PARL, POLR2H* | rs16858210 | 3 | 183624010 | 0.14 | G | A | 0.26 | 0.16 (0.07) | 5.62 | 0.24 | 0.14 (0.02) | 34.32 |
| *HELQ, FAM175A* | rs4693089 | 4 | 84373622 | 0.2 | A | G | 0.49 | 0.13 (0.06) | 4.42 | 0.49 | 0.29 (0.02) | 210.82 |
| *ASCL1, MLF1IP* | rs6856693 | 4 | 185748806 | 0.16 | A | G | 0.43 | 0.22 (0.06) | 12.45 | 0.42 | 0.23 (0.02) | 120.39 |
| *PAPD7* | rs427394 | 5 | 6745875 | 0.13 | G | A | 0.58 | 0.08 (0.06) | 1.58 | 0.58 | 0.13 (0.02) | 42.63 |
| *SH3PXD2B* | rs11738223 | 5 | 171934492 | 0.12 | A | G | 0.32 | 0.21 (0.07) | 10.46 | 0.33 | 0.08 (0.02) | 12.79 |
| *UIMC1* | rs2241584 | 5 | 175956177 | 0.14 | A | G | 0.62 | 0.21 (0.06) | 11.22 | 0.61 | 0.17 (0.02) | 62.94 |
| *UIMC1* | rs365132 | 5 | 176378574 | 0.24 | G | T | 0.49 | 0.36 (0.06) | 35.45 | 0.48 | 0.32 (0.02) | 239.44 |
| *SYCP2L, MAK* | rs6899676 | 6 | 10895260 | 0.23 | A | G | 0.2 | 0.23 (0.08) | 9.16 | 0.2 | 0.28 (0.03) | 121.07 |
| *SYCP2L, MAK* | rs9393800 | 6 | 10951737 | 0.17 | G | A | 0.73 | 0.13 (0.07) | 3.85 | 0.73 | 0.2 (0.02) | 77.18 |
| *MSH5, HLA* | rs2230365 | 6 | 31525448 | 0.17 | C | T | 0.15 | 0.33 (0.08) | 15.59 | 0.14 | 0.17 (0.03) | 33.93 |
| *MSH5, HLA* | rs707938 | 6 | 31729359 | 0.17 | G | A | 0.68 | 0.28 (0.06) | 18.79 | 0.68 | 0.18 (0.02) | 64.71 |
| *REV3L* | rs12196873 | 6 | 111598058 | 0.16 | A | C | 0.14 | 0.2 (0.09) | 5.64 | 0.14 | 0.15 (0.03) | 26.07 |
| *STAR* | rs2720044 | 8 | 37980587 | 0.29 | A | C | 0.14 | 0.16 (0.09) | 3.33 | 0.16 | 0.27 (0.03) | 98.66 |
| *CHD7* | rs10957156 | 8 | 61629401 | 0.14 | A | G | 0.23 | 0.15 (0.07) | 4.27 | 0.25 | 0.13 (0.02) | 29.68 |
| *APTX* | rs4879656 | 9 | 33012382 | 0.12 | A | C | 0.62 | 0.13 (0.06) | 4.11 | 0.62 | 0.11 (0.02) | 25.98 |
| *FBXO18* | rs10905065 | 10 | 5769827 | 0.11 | A | G | 0.39 | 0.06 (0.06) | 0.93 | 0.39 | 0.1 (0.02) | 22.21 |
| *FSHB* | rs11031006 | 11 | 30226528 | 0.22 | G | A | 0.14 | 0.25 (0.09) | 8.17 | 0.15 | 0.19 (0.03) | 44.73 |
| *LOC105376607* | rs6484478*,** | 11 | 30306440 | 0.1 | G | A | 0.21 | -0.1 (0.07) | 1.92 | 0.23 | -0.01 (0.02) | 0.16 |
| *EIF3M* | rs10734411 | 11 | 32541784 | 0.12 | A | G | 0.52 | 0.03 (0.06) | 0.31 | 0.52 | 0.13 (0.02) | 39.64 |
| *PRIM1, TAC3* | rs2277339 | 12 | 57146069 | 0.31 | G | T | 0.89 | 0.23 (0.1) | 5.13 | 0.89 | 0.4 (0.03) | 147.1 |
| *HELB* | rs3741604 | 12 | 66696410 | 0.09 | T | C | 0.45 | 0.05 (0.06) | 0.74 | 0.47 | 0.1 (0.02) | 24.14 |
| *HELB* | rs75770066 | 12 | 66704225 | 0.85 | A | G | 0.03 | 0.87 (0.21) | 16.64 | 0.03 | 1.05 (0.06) | 321.35 |
| *HELB* | rs1183272 | 12 | 66735421 | 0.07 | C | T | 0.58 | 0.17 (0.06) | 7.39 | 0.55 | 0.11 (0.02) | 28.69 |
| *HELB* | rs7397861* | 12 | 66814466 | 0.1 | G | C | 0.36 | 0 (0.06) | 0 | 0.37 | 0.07 (0.02) | 12.4 |
| *SPPL3, SRSF9* | rs551087** | 12 | 121209193 | 0.13 | G | A | 0.7 | 0.06 (0.07) | 0.72 | 0.71 | -0.01 (0.02) | 0.23 |
| *MPHOSPH9* | rs1727326 | 12 | 123600086 | 0.19 | C | G | 0.86 | 0.01 (0.1) | 0.01 | 0.86 | 0.17 (0.03) | 31.61 |
| *PIWIL1* | rs12824058 | 12 | 130804334 | 0.14 | G | A | 0.58 | 0.01 (0.06) | 0.01 | 0.58 | 0.12 (0.02) | 35.81 |
| *TDRD3* | rs4886238 | 13 | 61113739 | 0.18 | G | A | 0.33 | 0.31 (0.06) | 23.75 | 0.33 | 0.2 (0.02) | 86.62 |
| *APEX1, PARP2,PNP* | rs1713460 | 14 | 20933615 | 0.14 | G | A | 0.7 | 0.2 (0.07) | 9.15 | 0.69 | 0.15 (0.02) | 48.63 |
| *INO80, RAD51* | rs9796 | 15 | 41271447 | 0.13 | T | A | 0.54 | 0.14 (0.06) | 4.89 | 0.53 | 0.17 (0.02) | 71.01 |
| *POLG, FANCI* | rs1054875 | 15 | 89879126 | 0.19 | T | A | 0.58 | 0.02 (0.06) | 0.11 | 0.61 | 0.18 (0.02) | 78.23 |
| *C16orf72, ABAT* | rs9039 | 16 | 9205363 | 0.12 | C | T | 0.71 | 0.08 (0.07) | 1.48 | 0.73 | 0.13 (0.02) | 29.87 |
| *GSPT1, BCAR4* | rs10852344 | 16 | 12016919 | 0.16 | T | C | 0.42 | 0.14 (0.06) | 5.32 | 0.4 | 0.25 (0.02) | 147.01 |
| *UBE2MP1* | rs12599106 | 16 | 34498025 | 0.12 | A | T | 0.48 | 0.14 (0.06) | 4.68 | 0.5 | 0.13 (0.02) | 39.22 |
| *RPAIN* | rs8070740 | 17 | 5331896 | 0.15 | A | G | 0.25 | 0.09 (0.07) | 1.7 | 0.24 | 0.15 (0.02) | 38.01 |
| *STARD3,PGAP3, CDK12* | rs2941505 | 17 | 37832704 | 0.13 | A | G | 0.66 | 0.12 (0.06) | 3.61 | 0.69 | 0.16 (0.02) | 54.33 |
| *BRCA1* | rs1799949 | 17 | 41245466 | 0.14 | G | A | 0.33 | 0.14 (0.06) | 4.57 | 0.32 | 0.23 (0.02) | 109.02 |
| *ARID3A* | rs349306# | 19 | 950694 | 0.23 | G | A | 0.87 | 0.17 (0.1) | 2.74 | 0.87 | 0.24 (0.03) | 61.17 |
| *ZNF729* | rs7259376 | 19 | 22507705 | 0.11 | A | G | 0.55 | 0.24 (0.06) | 15.5 | 0.53 | 0.11 (0.02) | 30.08 |
| *BRSK1, NLRP11,U2AF2* | rs11668344 | 19 | 55833664 | 0.41 | G | A | 0.64 | 0.43 (0.06) | 46.77 | 0.63 | 0.49 (0.02) | 534.83 |
| *BRSK1,NLRP11,U2AF2* | rs2547274 | 19 | 56310228 | 0.28 | G | C | 0.09 | 0.27 (0.1) | 6.57 | 0.09 | 0.24 (0.04) | 44.53 |
| *BRSK1, NLRP11,U2AF2* | rs12461110 | 19 | 56320663 | 0.17 | A | G | 0.65 | 0.26 (0.06) | 16.32 | 0.63 | 0.17 (0.02) | 67.43 |
| *MCM8* | rs451417 | 20 | 5941999 | 0.2 | A | C | 0.88 | 0.05 (0.09) | 0.31 | 0.88 | 0.25 (0.03) | 66.81 |
| *MCM8* | rs16991615 | 20 | 5948227 | 0.88 | G | A | 0.06 | 0.85 (0.13) | 43.11 | 0.06 | 1.06 (0.04) | 623.76 |
| *SLCO4A1* | rs2236553# | 20 | 61289743 | 0.16 | C | T | 0.75 | 0.07 (0.08) | 0.78 | 0.75 | 0.22 (0.02) | 81.7 |
| *DIDO1* | rs13040088# | 20 | 61549202 | 0.16 | G | A | 0.8 | 0.2 (0.08) | 6.53 | 0.79 | 0.17 (0.02) | 46.8 |
| *CHEK2* | rs5762534 | 22 | 28633571 | 0.16 | T | C | 0.15 | 0.22 (0.08) | 7.19 | 0.15 | 0.13 (0.03) | 20.3 |
| *DMC1, DDX17* | rs763121 | 22 | 38879940 | 0.16 | G | A | 0.65 | 0.06 (0.06) | 0.92 | 0.66 | 0.17 (0.02) | 63.63 |

SNP = single nucleotide polymorphism; Chr = chromosome number; SE = standard error; WHI = Women's Health Initiative; UKB = UK Biobank. * indicates SNPs with an inconsistent direction of effect between Day and WHI, # indicates 3 SNPs that failed imputation on ~10% samples in WHI, ** indicates SNPs with an inconsistent direction of effect between Day and UKB. These SNPs were excluded from further analysis.

### Item 5

R-squared for imputation of SNPs in each data set. Women's Health Initiative (WHI) was comprised of 6 separate data sets whose R-squared values are each listed.

| SNP | rsID | ARIC | CHS | E2C2 | FHS | Kaiser | lung336 | lung634 | lung753 | MESA | NHS | UKB | WHI: GARNET | WHI: GECCO | WHI: HIPFX | WHI: MOPMAP | WHI: ONCOCHIP | WHI: WHIMS | WTCCC2 |
|---|---|---|---|---|---|---|---|---|---|---|---|---|---|---|---|---|---|---|---|
| 1:180962282 | rs1411478 | 0.6359 | 0.9974 | 0.9918 | 0.9873 | 0.9864 | 0.9999 | 0.9933 | 0.9983 | 0.6353 | 1.0001 | 1.0059 | 0.9947 | 0.9998 | 1.0000 | 0.9180 | 0.9564 | 0.9971 | 0.9993 |
| 1:242017826 | rs2236918 | 0.2832 | 0.7315 | 0.4130 | 0.9984 | 0.9832 | 0.8298 | 0.9854 | 0.7189 | 0.2033 | 0.8530 | 1.0020 | 0.9917 | 0.8952 | 0.8807 | 0.9787 | 0.9759 | 0.9915 | 0.8052 |
| 1:39380385 | rs4246511 | 0.3338 | 0.8366 | 0.8316 | 0.7318 | 0.9777 | 0.9117 | 0.9191 | 0.8541 | 0.3052 | 0.9060 | 0.9880 | 0.9990 | 0.9200 | 0.9257 | 0.9770 | 0.9985 | 0.9157 | 0.8944 |
| 1:46747301 | rs12142240 | 0.9099 | 0.9993 | 0.9995 | 0.8049 | 0.9837 | 1.0000 | 1.0000 | 0.9997 | 0.9154 | 1.0002 | 1.0092 | 0.9998 | 0.9999 | 1.0000 | 0.9862 | 0.9831 | 0.9998 | 1.0000 |
| 2:171941018 | rs930036 | 0.9541 | 0.9997 | 0.9999 | 0.9822 | 0.9893 | 0.9999 | 1.0000 | 1.0000 | 0.9533 | 0.9998 | 1.0104 | 1.0000 | 1.0000 | 0.9999 | 0.9887 | 1.0000 | 1.0000 | 0.9997 |
| 2:27716494 | rs704795 | 0.9419 | 0.9768 | 0.9780 | 0.9273 | 0.9938 | 0.9919 | 1.0000 | 0.9888 | 0.9494 | 0.9921 | 1.0068 | 1.0000 | 0.9924 | 0.9922 | 0.9805 | 0.9880 | 0.9998 | 0.9907 |
| 2:48018081 | rs1800932 | 0.9870 | 0.9906 | 0.9889 | 0.9988 | 0.9997 | 0.9940 | 0.9969 | 0.9919 | 0.9979 | 0.9856 | 1.0026 | 0.9996 | 0.9858 | 0.9874 | 0.9992 | 0.9846 | 0.9899 | 0.9924 |
| 3:183624010 | rs16858210 | 0.8833 | 0.9484 | 0.9791 | 0.9389 | 0.8778 | 0.9934 | 0.9985 | 0.9681 | 0.9055 | 0.9929 | 0.9948 | 0.9981 | 0.9885 | 0.9886 | 0.9514 | 0.9799 | 0.9983 | 0.9908 |
| 4:185748806 | rs6856693 | 0.9939 | 0.9218 | 0.9386 | 0.9961 | 0.9566 | 0.9581 | 0.9525 | 0.9387 | 0.9940 | 0.9048 | 1.0030 | 0.9996 | 0.9072 | 0.9067 | 0.7359 | 0.9476 | 0.9355 | 0.9465 |
| 4:84373622 | rs4693089 | 0.9921 | 0.9961 | 0.8266 | 0.9893 | 0.9995 | 0.9991 | 0.9993 | 0.9992 | 0.9952 | 0.9992 | 1.0096 | 0.9998 | 0.9996 | 0.9997 | 0.9999 | 0.9876 | 0.9997 | 0.9992 |
| 5:171934492 | rs11738223 | 0.8475 | 0.9640 | 0.9517 | 0.9693 | 0.9604 | 0.9774 | 0.9598 | 0.9694 | 0.8583 | 0.9731 | 1.0007 | 0.9625 | 0.9713 | 0.9723 | 0.9513 | 0.9593 | 0.9599 | 0.9776 |
| 5:175956177 | rs2241584 | 0.9985 | 0.9987 | 0.8420 | 0.9979 | 0.6030 | 0.9993 | 0.9859 | 0.9993 | 0.9990 | 0.9998 | 0.9763 | 0.9913 | 0.9996 | 0.9995 | 0.4519 | 0.9549 | 0.9855 | 0.9995 |
| 5:176378574 | rs365132 | 0.9189 | 0.9991 | 0.9990 | 0.9543 | 0.9999 | 0.9999 | 1.0000 | 0.9999 | 0.8932 | 1.0002 | 1.0071 | 1.0000 | 1.0000 | 1.0000 | 0.9732 | 0.9872 | 1.0000 | 1.0000 |
| 5:6745875 | rs427394 | 0.9999 | 0.9654 | 0.9581 | 1.0000 | 0.9946 | 0.9971 | 0.9971 | 0.9656 | 1.0000 | 0.9960 | 0.9998 | 0.9971 | 0.9965 | 0.9967 | 0.9953 | 0.8465 | 0.9938 | 0.9933 |
| 6:10895260 | rs6899676 | 0.9430 | 0.9437 | 0.9820 | 0.9586 | 0.9841 | 0.9907 | 0.9901 | 0.9706 | 0.9420 | 0.9893 | 0.9889 | 0.9844 | 0.9870 | 0.9814 | 0.9870 | 0.9995 | 0.9879 | 0.9855 |
| 6:10951737 | rs9393800 | 0.9432 | 0.9146 | 0.7457 | 0.9089 | 0.9712 | 0.9770 | 0.9882 | 0.9476 | 0.9658 | 0.9839 | 0.9747 | 0.9808 | 0.9756 | 0.9712 | 0.9948 | 0.9825 | 0.9910 | 0.9724 |
| 6:111598058 | rs12196873 | 0.9998 | 0.9515 | 0.9590 | 0.9996 | 0.9825 | 0.9750 | 0.9794 | 0.9723 | 1.0002 | 0.9664 | 1.0041 | 0.9984 | 0.9678 | 0.9745 | 0.9765 | 0.9827 | 0.9859 | 0.9787 |
| 6:31525448 | rs2230365 | 0.9060 | 0.9950 | 0.9961 | 0.9064 | 0.9740 | 0.9988 | 0.9986 | 0.9959 | 0.9219 | 0.9988 | 1.0041 | 0.9984 | 0.9981 | 0.9971 | 0.9158 | 0.9989 | 0.9992 | 0.9988 |
| 6:31729359 | rs707938 | 0.8196 | 0.9990 | 0.9756 | 0.9355 | 0.9883 | 0.9911 | 0.9996 | 0.9799 | 0.7668 | 0.9937 | 1.0093 | 0.9996 | 0.9909 | 0.9919 | 0.9955 | 0.9999 | 0.9995 | 0.9802 |
| 8:37980587 | rs2720044 | 0.9896 | 0.7201 | 0.7297 | 0.9967 | 0.8295 | 0.8309 | 0.6339 | 0.7350 | 0.9865 | 0.7505 | 0.9941 | 0.9842 | 0.7437 | 0.7607 | 0.8343 | 0.9987 | 0.8530 | 0.7463 |
| 8:61629401 | rs10957156 | 0.9845 | 0.9562 | 0.9817 | 0.9966 | 0.9877 | 0.9955 | 0.9931 | 0.9899 | 0.9843 | 0.9922 | 1.0001 | 0.9918 | 0.9877 | 0.9891 | 0.9901 | 0.9334 | 0.9904 | 0.9938 |
| 9:33012382 | rs4879656 | 0.9382 | 0.9866 | 0.9862 | 0.9623 | 0.9531 | 1.0000 | 0.9948 | 0.9927 | 0.7439 | 0.9998 | 1.0049 | 0.9946 | 1.0000 | 1.0000 | 0.9910 | 0.9621 | 0.9963 | 0.9995 |
| 10:5769827 | rs10905065 | 0.9976 | 0.9914 | 0.9861 | 0.9980 | 0.9936 | 0.9963 | 0.9878 | 0.9946 | 0.9978 | 0.9950 | 1.0047 | 0.9932 | 0.9961 | 0.9960 | 0.9943 | 0.9773 | 0.9927 | 0.9955 |
| 11:30226528 | rs11031006 | 0.9958 | 0.9154 | 0.9981 | 0.9924 | 0.9098 | 0.9999 | 0.9995 | 0.9535 | 0.9981 | 1.0001 | 0.9932 | 0.9997 | 0.9999 | 0.9999 | 0.9212 | 0.9701 | 0.9999 | 0.9992 |
| 11:30306440 | rs6484478 | 0.9924 | 0.9928 | 0.9936 | 0.9922 | 0.9900 | 0.9971 | 0.9988 | 0.9978 | 0.9935 | 0.9980 | 1.0060 | 1.0000 | 0.9962 | 0.9977 | 0.9949 | 0.9055 | 0.9989 | 0.9984 |
| 11:32541784 | rs10734411 | 0.9992 | 0.9372 | 0.8689 | 0.9990 | 0.9993 | 0.9781 | 0.9868 | 0.9745 | 0.9997 | 0.9783 | 1.0131 | 0.9432 | 0.9788 | 0.9770 | 0.9996 | 0.9585 | 0.8401 | 0.9777 |
| 12:121209193 | rs551087 | 0.9645 | 0.9919 | 0.9795 | 0.9677 | 0.9958 | 0.9891 | 0.9918 | 0.9958 | 0.9647 | 0.9970 | 1.0086 | 0.9953 | 0.9706 | 0.9976 | 0.9967 | 0.9528 | 0.9948 | 0.9969 |
| 12:123600086 | rs1727326 | 0.7142 | 0.7745 | 0.7080 | 0.6900 | 0.7844 | 0.8793 | 0.8352 | 0.8266 | 0.6776 | 0.7698 | 0.9900 | 0.7721 | 0.7696 | 0.7887 | 0.5361 | 0.7008 | 0.7439 | 0.8740 |
| 12:130804334 | rs12824058 | 0.4968 | 0.9574 | 0.9211 | 0.8772 | 0.9997 | 0.9878 | 0.9807 | 0.9339 | 0.4956 | 0.9873 | 1.0006 | 0.9876 | 0.9879 | 0.9869 | 0.9996 | 0.9478 | 0.9909 | 0.9746 |
| 12:57146069 | rs2277339 | 0.2765 | 0.9976 | 0.9983 | 0.4193 | 0.9991 | 0.9996 | 0.9985 | 0.9972 | 0.3172 | 0.9957 | 1.0021 | 0.9963 | 0.9969 | 0.9962 | 0.9887 | 0.9924 | 0.9948 | 0.9991 |
| 12:66696410 | rs3741604 | 0.9761 | 0.9945 | 0.9736 | 0.9863 | 0.9654 | 0.9995 | 0.9999 | 0.9999 | 0.9765 | 1.0001 | 1.0076 | 1.0000 | 0.9895 | 1.0000 | 0.9398 | 0.9691 | 0.9941 | 0.9995 |
| 12:66704225 | rs75770066 | 0.2234 | 0.3721 | 0.6198 | 0.1991 | 0.8239 | 0.6927 | 0.6368 | 0.4224 | 0.2535 | 0.4072 | 1.0034 | 0.5468 | 0.2571 | 0.4710 | 0.4832 | 0.7880 | 0.4696 | 0.6156 |
| 12:66735421 | rs1183272 | 0.9922 | 0.9757 | 0.9350 | 0.9887 | 0.9967 | 0.9958 | 0.9949 | 0.9936 | 0.9896 | 0.9938 | 1.0014 | 0.9975 | 0.9986 | 0.9924 | 0.9969 | 0.9705 | 0.9958 | 0.9972 |
| 12:66814466 | rs7397861 | 0.5640 | 0.9219 | 0.9930 | 0.8642 | 0.9911 | 0.9940 | 0.9976 | 0.9837 | 0.6006 | 0.9949 | 1.0224 | 0.9981 | 0.9968 | 0.9927 | 0.9922 | 0.9612 | 0.9988 | 0.9939 |
| 13:61113739 | rs4886238 | 0.6103 | 0.9623 | 0.9763 | 0.9222 | 0.9732 | 0.9946 | 0.9797 | 0.9810 | 0.5914 | 0.9912 | 0.9962 | 0.9884 | 0.9896 | 0.9895 | 0.9840 | 0.9675 | 0.9880 | 0.9949 |
| 14:20933615 | rs1713460 | 0.2353 | 0.6526 | 0.7824 | 0.2441 | 0.4436 | 0.7006 | 0.7856 | 0.7940 | 0.2596 | 0.9844 | 1.0031 | 0.9960 | 0.9828 | 0.9869 | 0.7744 | 0.9956 | 0.9934 | 0.7978 |
| 15:41271447 | rs9796 | 0.6573 | 0.8547 | 0.9337 | 0.8227 | 0.8005 | 0.9385 | 0.9577 | 0.9356 | 0.6428 | 0.9525 | 1.0034 | 0.9673 | 0.9443 | 0.9474 | 0.8701 | 0.8381 | 0.9639 | 0.9475 |
| 15:89879126 | rs1054875 | 0.8816 | 0.9337 | 0.9704 | 0.9396 | 0.5590 | 0.9744 | 0.9786 | 0.9673 | 0.9273 | 0.9761 | 0.9963 | 0.9736 | 0.5884 | 0.9765 | 0.6900 | 0.9553 | 0.9776 | 0.9744 |
| 16:12016919 | rs10852344 | 0.7901 | 0.8137 | 0.9996 | 0.7803 | 0.8858 | 0.9997 | 0.9998 | 0.9993 | 0.8004 | 0.9998 | 1.0200 | 0.9993 | 0.9997 | 0.8217 | 0.8353 | 0.9980 | 0.9993 | 0.9997 |
| 16:34498025 | rs12599106 | 0.1149 | 0.3292 | 0.2424 | 0.5909 | 0.7164 | 0.6434 | 0.5288 | 0.3209 | 0.1166 | 0.8905 | 1.0168 | 0.9416 | 0.7841 | 0.7988 | 0.9524 | 0.5968 | 0.8776 | 0.3412 |
| 16:9205363 | rs9039 | 0.9904 | 0.9992 | 0.9148 | 0.9997 | 0.9186 | 0.9998 | 0.9666 | 0.9997 | 0.9867 | 0.9996 | 0.9938 | 0.9925 | 0.9776 | 0.9998 | 0.9680 | 0.9710 | 0.9930 | 0.9995 |
| 17:37832704 | rs2941505 | 0.9945 | 0.9534 | 0.9718 | 0.9877 | 0.9247 | 0.9976 | 0.9995 | 0.9797 | 0.9961 | 0.9971 | 1.0042 | 0.9989 | 0.9981 | 0.9642 | 0.9532 | 0.9790 | 0.9988 | 0.9974 |
| 17:41245466 | rs1799949 | 0.8906 | 0.9250 | 0.9305 | 0.8997 | 0.9988 | 0.9952 | 0.9241 | 0.9993 | 0.6984 | 0.9997 | 1.0039 | 0.9999 | 0.9991 | 0.9996 | 0.9996 | 0.9333 | 0.9998 | 0.9137 |
| 17:5331896 | rs8070740 | 0.9427 | 0.9376 | 0.9060 | 0.9422 | 0.9901 | 0.9701 | 0.9846 | 0.9641 | 0.9219 | 0.9588 | 1.0142 | 0.9955 | 0.9646 | 0.9659 | 0.9914 | 0.9767 | 0.9898 | 0.9698 |
| 19:22507705 | rs7259376 | 0.9994 | 0.9555 | 0.9791 | 0.9993 | 0.9522 | 0.9795 | 0.9989 | 0.9922 | 0.9999 | 0.9960 | 1.0032 | 0.9981 | 0.9861 | 0.9893 | 0.9059 | 0.8767 | 0.9988 | 0.9919 |
| 19:55833664 | rs11668344 | 0.9579 | 0.9654 | 0.9660 | 0.7871 | 0.9571 | 0.9949 | 0.9973 | 0.9867 | 0.9574 | 0.9944 | 1.0036 | 0.9974 | 0.9825 | 0.9945 | 0.8730 | 0.9998 | 0.9980 | 0.9947 |
| 19:56310228 | rs2547274 | 0.4737 | 0.8493 | 0.8722 | 0.5976 | 0.9873 | 0.9063 | 0.9715 | 0.8823 | 0.5000 | 0.8464 | 0.9722 | 0.9698 | 0.4695 | 0.9118 | 0.9047 | 0.8598 | 0.9838 | 0.8976 |
| 19:56320663 | rs12461110 | 0.9960 | 0.9973 | 0.9973 | 0.7854 | 0.9966 | 0.9954 | 0.9965 | 0.9969 | 0.9959 | 0.9883 | 1.0096 | 0.9915 | 0.5908 | 0.9793 | 0.9889 | 0.9872 | 0.9967 | 0.9952 |
| 19:950694 | rs349306 | 0.1631 | 0.7395 | 0.7822 | 0.1301 | 0.0000 | 0.8489 | 0.8775 | 0.7637 | 0.1750 | 0.7363 | 0.9663 | 0.9059 | 0.7416 | 0.7019 | | 0.7935 | 0.8607 | 0.7939 |
| 20:5941999 | rs451417 | 0.2533 | 0.9014 | 0.9045 | 0.9125 | 0.9925 | 0.9842 | 0.9938 | 0.9595 | 0.2568 | 0.9756 | 1.0315 | 0.9855 | 0.9811 | 0.9736 | 0.9721 | 0.9813 | 0.9818 | 0.9733 |
| 20:5948227 | rs16991615 | 0.9616 | 0.7064 | 0.7602 | 0.6089 | 0.9953 | 0.9968 | 0.7877 | 0.7151 | 0.9949 | 0.9931 | 1.0004 | 0.9892 | 0.9960 | 0.9942 | 0.9890 | 0.9931 | 0.7854 | 0.9942 |
| 20:61289743 | rs2236553 | 0.9185 | 0.6193 | 0.8636 | 0.3387 | 0.0115 | 0.9912 | 0.9124 | 0.9867 | 0.9215 | 0.9910 | 0.9900 | 0.9956 | | 0.8953 | 0.1658 | | 0.9364 | 0.9943 |
| 20:61549202 | rs13040088 | 0.9972 | 0.9657 | 0.9512 | 0.8900 | 0.8672 | 0.9909 | 0.9801 | 0.9818 | 0.9975 | 0.9891 | 0.9988 | 0.9956 | | 0.9875 | 0.9235 | | 0.9953 | 0.9869 |
| 22:28633571 | rs5762534 | 0.8612 | 0.9253 | 0.9996 | 0.9223 | 0.9996 | 0.9999 | 0.9999 | 0.9745 | 0.9029 | 0.9998 | 1.0006 | 0.9993 | 0.9989 | 0.9993 | 0.9995 | 0.9997 | 0.9997 | 0.9996 |
| 22:38879940 | rs763121 | 0.8313 | 0.8320 | 0.9230 | 0.6462 | 0.7968 | 0.7501 | 0.9999 | 0.8868 | 0.9643 | 0.7207 | 0.9996 | 0.9998 | 0.9467 | 0.9510 | 0.9259 | 0.9142 | 1.0000 | 0.9416 |

#### Item 6

##### Details on genetic ancestry:

Participants in WHI and UKB were of European genetic ancestry. This ancestry had already been determined by the respective datasets using principal components (PCs), and thus we did not additionally adjust for these PCs.

##### WHI:

The study population consisted of all 161,808 post-menopausal women enrolled in the WHI clinical trials (WHI-CT) or observational study (WHI-OS) between 1993 and 1998 with follow up through 2015. Women with history of bilateral oophorectomy before menopausal symptoms were excluded. Only women who reported European American ancestry were included given the genetic instruments to be used for ANM were derived from European populations. Further exclusions were variable and depended on the availability of covariates, the outcome or quantitative trait being examined (n = 8,740). For most observational analyses, these exclusions resulted in a sample size of ~98,000.

Genome-wide genotyping data were available in a subset of 15,136 participants with European American ancestry. These genetic data were generated over the last approximately 9 years through ancillary and core sub studies in WHI. In 2014, WHI investigators at the Data Coordinating Center harmonized all genome-wide data generated to date through imputation to 1000 genomes reference panel. Additional details for this harmonization process can be found online at the database of Genotypes and Phenotypes (WHI sub study phs000746; https://www.ncbi.nlm.nih.gov/projects/gap/cgi-bin/study.cgi?study_id=phs000746.v3.p3). The harmonization process produced a variable called InconsistentEthnicity which represented those whose self-reported race and ancestry did not match. We found that only 3 subjects in our dataset had the InconsistentEthnicity value true. We additionally included another 8789 subjects with European ancestry which have genome-wide genotyping data from Oncochip and imputed to 1000 Genomes reference panel. Applying all the exclusion criteria described above resulted in a sample size of ~17,900 for most IV analyses.

##### UKB:

The study population consisted of 154,626 women who had responded to question #3581, the age of menopause, with a positive numeric value. After exclusion of those who had undergone bilateral oophorectomy or hysterectomy prior to 2 years after the age provided, the data consisted of 141,013 women who had undergone natural menopause. In 2018, UK Biobank investigators performed sample quality control including determination of genetic ethnic grouping (variable 22006) and relatedness (variable 22021) as described at https://biobank.ctsu.ox.ac.uk/crystal/ukb/docs/ukb_genetic_data_description.txt. After exclusion of those of non-European ancestry and individuals with third-degree or closer relatives, the sample size was up to 95,469 for each IV analysis.

##### Other data sets:

These data were primarily of European ancestry. Exceptions were the Atherosclerosis Risk in Communities Study (ARIC), the Multi-Ethnic Study of Atherosclerosis (MESA), and the Kaiser Permanente Research Program on Genes, Environment, and Health, which we limited to those of likely European ancestry. Ancestry, race, or ethnicity were reported, and these data were often self-reported.
Descriptions of the data from the NIH database of Genotypes and Phenotypes (dbGaP) were as follows:
**Nurses Health Study I**: Reported in dbGaP as European descent.
**E2C2**: Reported in dbGaP as European descent.
**PLCO + CSP-II (phs000336)**: Reported in dbGaP as European descent.
**NCI GWAS in never smokers (phs000634)**: Reported in dbGaP as Caucasian and Whites.
**High density SNP analysis (phs000753)**: Reported in dbGaP as European ancestry.
**ARIC**: Extracted from the “racegrp” variable with options African American and White.

**CHS**: Extracted from the “RACE01” variable. The vast majority in the original study were White; other options were Black, American Indian/Alaskan Native, Asian/Pacific Islander, and Other.
**FHS**: Extracted participants marked “Caucasian”.
**MESA**: Extracted from the “race1c” variable. Options were White/Caucasian, Chinese American, Black/African American, and Hispanic.
**WTCCC2**: This data was extracted from the European Genome-Phenome Archive and was from a British population.
**Kaiser**: Participants self-identified as African American, Asian, Latino, or Non-Hispanic White. A different genetic array was then designed for each race-ethnic group.

### Supplemental Results

#### Item 7

Table. Observational association results. Odds ratio (OR) and 95% confidence interval (CI).

| Outcome and dataset | OR [95% CI] | Cases | Total |
|---|---|---|---|
| Breast cancer | | | |
| WHI | 1.06 [1.03, 1.08] | 8087 | 97703 |
| UKB | 1.15 [1.11, 1.19] | 3192 | 91205 |
| WHI + UKB summary | 1.10 [1.01, 1.19] | 11279 | 188908 |
| Endometrial cancer | | | |
| WHI | 1.20 [1.12, 1.27] | 1322 | 69553 |
| UKB | 1.37 [1.27, 1.48] | 748 | 95151 |
| WHI + UKB summary | 1.28 [1.12, 1.46] | 2070 | 164704 |
| Ovarian cancer | | | |
| WHI | 1.07 [1.00, 1.13] | 899 | 92111 |
| UKB | 1.03 [0.94, 1.12] | 419 | 95304 |
| WHI + UKB summary | 1.05 [1.00, 1.11] | 1318 | 187415 |
| Lung cancer | | | |
| WHI | 0.92 [0.89, 0.96] | 2509 | 98113 |
| UKB | 0.85 [0.81, 0.90] | 815 | 95390 |
| WHI + UKB summary | 0.89 [0.83, 0.96] | 3324 | 193503 |
| Coronary heart disease | | | |
| WHI | 0.97 [0.95, 0.99] | 4583 | 94024 |
| UKB | 0.95 [0.91, 0.99] | 1798 | 94074 |
| WHI + UKB summary | 0.97 [0.95, 0.98] | 6381 | 188098 |
| Ischemic stroke | | | |
| WHI | 0.96 [0.93, 0.99] | 2726 | 96969 |
| UKB | 0.92 [0.87, 0.96] | 1171 | 95273 |
| WHI + UKB summary | 0.94 [0.90, 0.99] | 3897 | 192242 |
| Fracture | | | |
| WHI | 0.96 [0.94, 0.99] | 8055 | 98112 |
| UKB | 0.87 [0.80, 0.94] | 481 | 95258 |
| WHI + UKB summary | 0.92 [0.83, 1.02] | 8536 | 193370 |
| Osteoporosis | | | |
| WHI | 0.95 [0.93, 0.96] | 25477 | 97726 |
| UKB | 0.84 [0.82, 0.87] | 4962 | 91547 |
| WHI + UKB summary | 0.89 [0.80, 1.00] | 30439 | 189273 |
| Alzheimer disease | | | |
| WHI | 0.94 [0.91, 0.97] | 3611 | 73798 |
| UKB | 0.89 [0.83, 0.95] | 611 | 95462 |
| WHI + UKB summary | 0.92 [0.88, 0.97] | 4222 | 169260 |

#### Item 8

Associations of ANM and confounders with instrument variables. ANM: Age of natural menopause, BMI: body mass index, MHT: menopausal hormone therapy, SBP: systolic blood pressure, TIA: trans ischemic attack, UKB: UK Biobank, WHI: Women's Health Initiative, WHO: World Health Organization.

**Supplementary Figure (a) for Item 8.**

**Units: ANM and age – years of age; BMI – kg/m$^2$; SBP – mmHg; education and income – ordinal categories; alcohol – servings/week; physical activity – kcal/week/kg; dietary vitamin D – mcg; serum vitamin D – nmol/L.**

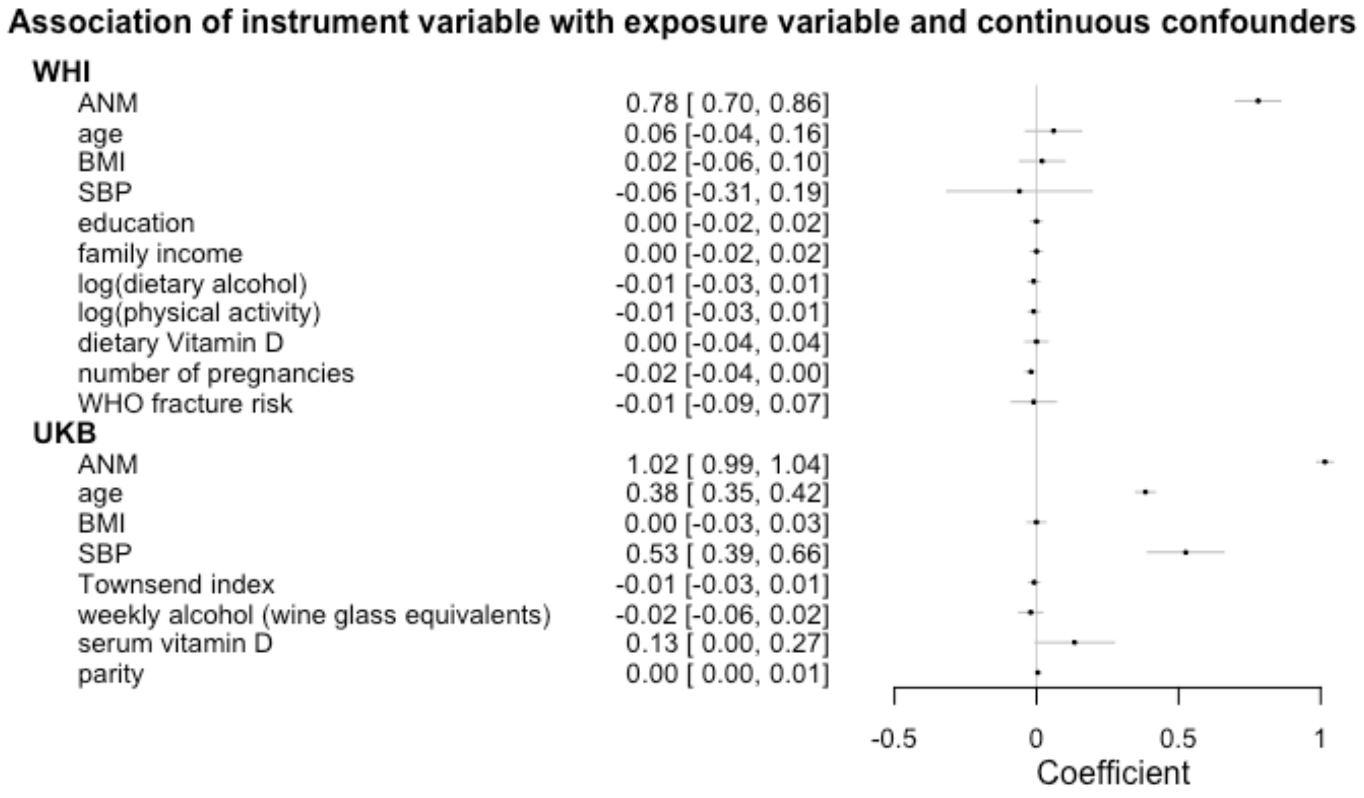

**Supplementary Figure (b) for Item 8.**

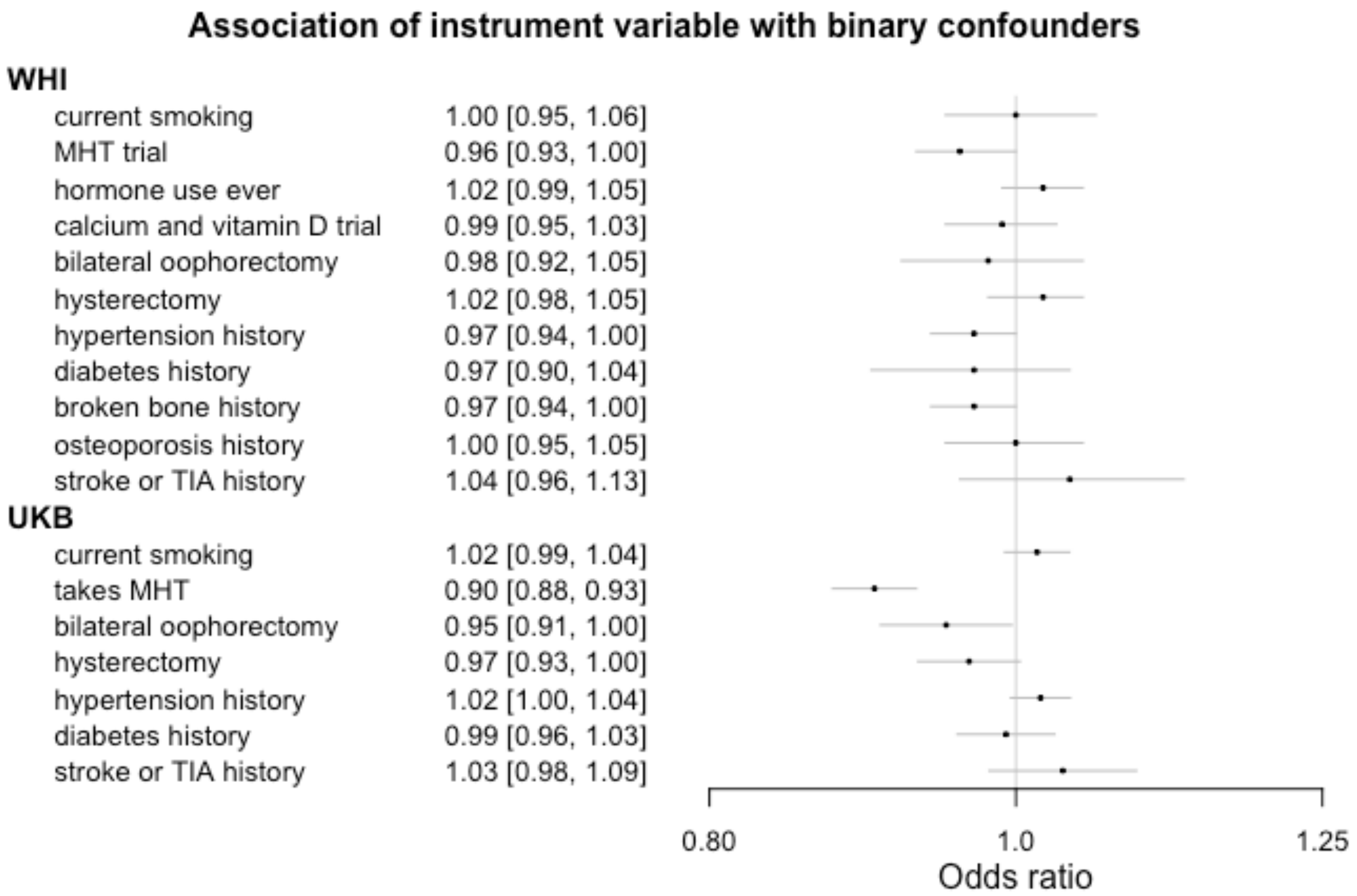

#### Item 9.

| Table. Mendelian Randomization association results by MR method. Odds ratio [confidence interval]. | | | | | |
|---|---|---|---|---|---|
| **Outcome and datasets** | **IVW** | **Weighted Median** | **MR-Egger** | **Cases** | **Total** |
| Breast cancer | | | | | |
| WHI | 1.82 [1.45, 2.28] | 2.05 [1.50, 2.80] | 1.92 [1.29, 2.85] | 3967 | 17906 |
| UKB | 0.98 [0.79, 1.22] | 1.06 [0.79, 1.43] | 1.14 [0.74, 1.74] | 3192 | 91205 |
| NHS | 1.57 [0.94, 2.62] | 1.79 [0.85, 3.76] | 2.35 [0.95, 5.79] | 1089 | 2182 |
| Summary for all data | 1.39 [0.88, 2.20] | 1.54 [0.93, 2.53] | 1.61 [1.05, 2.45] | 8248 | 111293 |
| Endometrial cancer | | | | | |
| WHI | 2.37 [1.18, 4.76] | 2.75 [0.94, 8.06] | 2.01 [0.61, 6.59] | 208 | 13200 |
| UKB | 1.37 [0.96, 1.96] | 1.38 [0.77, 2.48] | 1.76 [0.89, 3.48] | 748 | 95151 |
| E2C2 | 0.89 [0.58, 1.36] | 0.90 [0.47, 1.70] | 0.49 [0.24, 1.01] | 1830 | 3441 |
| Summary for all data | 1.34 [0.83, 2.15] | 1.33 [0.79, 2.26] | 1.15 [0.45, 2.93] | 2786 | 111792 |
| Ovarian cancer | | | | | |
| WHI | 1.19 [0.41, 3.49] | 0.81 [0.19, 3.38] | 0.66 [0.10, 4.36] | 134 | 17059 |
| UKB | 1.40 [0.86, 2.28] | 0.97 [0.46, 2.03] | 1.12 [0.43, 2.93] | 419 | 95304 |
| Summary for all data | 1.36 [0.87, 2.12] | 0.93 [0.48, 1.80] | 1.01 [0.43, 2.37] | 553 | 112363 |
| Lung cancer | | | | | |
| WHI | 1.52 [0.96, 2.41] | 1.78 [0.90, 3.52] | 1.51 [0.69, 3.32] | 558 | 17981 |
| UKB | 1.34 [0.95, 1.90] | 1.16 [0.68, 1.98] | 1.57 [0.80, 3.09] | 815 | 95390 |
| High density SNP analysis | 1.45 [0.54, 3.91] | 2.20 [0.54, 8.96] | 1.25 [0.21, 7.48] | 498 | 986 |
| NCI GWAS | 1.74 [0.75, 4.06] | 1.73 [0.59, 5.09] | 2.45 [0.56, 10.74] | 369 | 741 |
| PLCO+CSP-II GWAS | 0.89 [0.47, 1.70] | 0.54 [0.21, 1.37] | 0.70 [0.23, 2.10] | 835 | 1285 |
| Summary for all data | 1.35 [1.06, 1.71] | 1.27 [0.83, 1.93] | 1.41 [0.91, 2.16] | 3075 | 116383 |
| Coronary heart disease | | | | | |
| WHI | 0.83 [0.61, 1.13] | 0.77 [0.49, 1.20] | 0.67 [0.39, 1.16] | 1348 | 17357 |
| UKB | 0.94 [0.74, 1.19] | 0.81 [0.56, 1.19] | 0.85 [0.53, 1.36] | 1798 | 94074 |
| ARIC | 1.00 [0.55, 1.81] | 1.45 [0.59, 3.53] | 2.41 [0.91, 6.38] | 468 | 1902 |
| CHS | 1.35 [0.73, 2.51] | 2.15 [0.87, 5.30] | 1.77 [0.60, 5.27] | 495 | 4872 |
| FHS | 0.55 [0.16, 1.87] | 0.30 [0.05, 1.94] | 0.05 [0.01, 0.49] | 118 | 1294 |
| MESA | 2.20 [0.42, 11.64] | 1.86 [0.18, 19.66] | 0.79 [0.04, 13.90] | 66 | 1089 |
| Summary for all data | 0.93 [0.79, 1.11] | 0.96 [0.67, 1.38] | 0.91 [0.49, 1.70] | 4293 | 120588 |
| Ischemic stroke | | | | | |
| WHI | 0.58 [0.37, 0.92] | 0.57 [0.31, 1.05] | 0.50 [0.23, 1.10] | 830 | 17723 |
| UKB | 1.27 [0.93, 1.73] | 1.03 [0.66, 1.60] | 0.93 [0.50, 1.72] | 1171 | 95273 |
| ARIC | 0.60 [0.22, 1.62] | 0.35 [0.08, 1.53] | 0.57 [0.10, 3.28] | 220 | 1925 |
| CHS | 1.39 [0.60, 3.25] | 2.81 [0.78, 10.15] | 1.71 [0.38, 7.65] | 157 | 4878 |
| FHS | 0.60 [0.09, 4.03] | 0.65 [0.04, 9.69] | 1.67 [0.06, 49.06] | 1593 | 3193 |
| MESA | 0.87 [0.09, 8.41] | 0.53 [0.02, 14.42] | 13.20 [0.30, 573.80] | 45 | 1303 |
| WTCCC2 | 0.99 [0.48, 2.05] | 1.33 [0.69, 2.57] | 1.64 [0.47, 5.71] | 32 | 1089 |
| Summary for all data | 0.92 [0.65, 1.29] | 0.93 [0.62, 1.41] | 0.89 [0.58, 1.35] | 4048 | 125384 |
| Fracture | | | | | |
| WHI | 0.72 [0.58, 0.90] | 0.65 [0.47, 0.90] | 0.59 [0.41, 0.85] | 3310 | 17981 |
| UKB | 0.94 [0.60, 1.47] | 1.04 [0.53, 2.04] | 1.67 [0.69, 4.01] | 481 | 95258 |
| Summary for all data | 0.76 [0.62, 0.94] | 0.75 [0.49, 1.13] | 0.92 [0.33, 2.50] | 3791 | 113239 |
| Osteoporosis | | | | | |
| WHI | 0.95 [0.76, 1.18] | 0.96 [0.73, 1.27] | 1.29 [0.89, 1.87] | 4965 | 17960 |
| UKB | 0.74 [0.64, 0.86] | 0.75 [0.61, 0.94] | 0.84 [0.63, 1.12] | 4962 | 91547 |
| Kaiser | 0.76 [0.63, 0.93] | 0.72 [0.55, 0.94] | 0.86 [0.62, 1.19] | 5147 | 27573 |
| Summary for all data | 0.80 [0.70, 0.92] | 0.80 [0.68, 0.94] | 0.96 [0.74, 1.24] | 15074 | 137080 |
| Alzheimer disease | | | | | |
| WHI | 1.06 [0.68, 1.65] | 1.42 [0.79, 2.54] | 1.03 [0.47, 2.25] | 933 | 14740 |
| UKB | 0.97 [0.65, 1.45] | 0.99 [0.55, 1.79] | 1.48 [0.68, 3.21] | 611 | 95462 |
| Summary for all data | 1.01 [0.75, 1.36] | 1.19 [0.79, 1.80] | 1.24 [0.71, 2.14] | 1544 | 110202 |

Item 10.

Table 10a. Estimate, standard error, and p value from IVW MR analysis.

| Outcome | Estimate | Standard error | P-value |
|---|---|---|---|
| **Breast cancer** | | | |
| WHI | 0.5988 | 0.1150 | 1.90E-07 |
| UKB | -0.0203 | 0.1102 | 0.8537 |
| NHS | 0.4513 | 0.2619 | 0.0848 |
| **Summary for all data** | **0.3317** | **0.2340** | **0.1564** |
| **Endometrial cancer** | | | |
| WHI | 0.8629 | 0.3558 | 0.0153 |
| UKB | 0.3155 | 0.1835 | 0.0855 |
| E2C2 | -0.1202 | 0.2184 | 0.5821 |
| **Summary for all data** | **0.2908** | **0.2411** | **0.2278** |
| **Ovarian cancer** | | | |
| WHI | 0.1740 | 0.5490 | 0.7513 |
| UKB | 0.3369 | 0.2490 | 0.1760 |
| **Summary for all data** | **0.3091** | **0.2267** | **0.1728** |
| **Lung cancer** | | | |
| WHI | 0.4187 | 0.2352 | 0.0750 |
| UKB | 0.2954 | 0.1775 | 0.0961 |
| PLCO+CSP-II GWAS | -0.1166 | 0.3304 | 0.7241 |
| High density SNP analysis | 0.3702 | 0.5063 | 0.4647 |
| NCI GWAS | 0.5547 | 0.4318 | 0.1989 |
| **Summary for all data** | **0.2974** | **0.1211** | **0.0140** |
| **Coronary heart disease** | | | |
| WHI | -0.1863 | 0.1574 | 0.2366 |
| UKB | -0.0616 | 0.1223 | 0.6147 |
| ARIC | 0.0034 | 0.3023 | 0.9909 |
| CHS | 0.3022 | 0.3162 | 0.3392 |
| FHS | -0.6004 | 0.6246 | 0.3364 |
| MESA | 0.7888 | 0.8497 | 0.3532 |
| **Summary for all data** | **-0.0683** | **0.0870** | **0.4325** |
| **Ischemic stroke** | | | |
| WHI | -0.5447 | 0.2354 | 0.0207 |
| UKB | 0.2368 | 0.1597 | 0.1382 |
| ARIC | -0.5042 | 0.5042 | 0.3173 |
| CHS | 0.3294 | 0.4330 | 0.4468 |
| FHS | -0.5139 | 0.9729 | 0.5973 |
| MESA | -0.1362 | 1.1558 | 0.9062 |
| WTCCC2 | -0.0072 | 0.3710 | 0.9845 |
| **Summary for all data** | **-0.0865** | **0.1760** | **0.6231** |
| **Fracture** | | | |
| WHI | -0.3285 | 0.1138 | 3.91E-03 |
| UKB | -0.0661 | 0.2303 | 0.7741 |
| **Summary for all data** | **-0.2737** | **0.1067** | **0.0103** |
| **Osteoporosis** | | | |
| WHI | -0.0513 | 0.1106 | 0.6429 |
| UKB | -0.2965 | 0.0744 | 6.78E-05 |
| Kaiser | -0.2687 | 0.0976 | 5.92E-03 |
| **Summary for all data** | **-0.2217** | **0.0716** | **1.96E-03** |
| **Alzheimer disease** | | | |
| WHI | 0.0583 | 0.2258 | 0.7963 |
| UKB | -0.0313 | 0.2050 | 0.8785 |
| **Summary for all data** | **0.0092** | **0.1518** | **0.9519** |

Table 10b. False discovery rate calculation for multiple testing from IVW MR analysis. From online calculator https://tools.carbocation.com/FDR.

| Rank | ID | Original P value | Critical Value | Benjamini-Hochberg Adjusted P value | Significant using an FDR of 0.05? |
|---|---|---|---|---|---|
| 1 | Osteoporosis | 0.001957565 | 0.005555556 | 0.017618085 | Yes |
| 2 | Fracture | 0.010297127 | 0.011111111 | 0.046337072 | Yes |
| 3 | Lung | 0.014009885 | 0.016666667 | 0.042029655 | Yes |
| 4 | Breast cancer | 0.156427737 | 0.022222222 | 0.351962408 | No |
| 5 | Ovarian | 0.172817107 | 0.027777778 | 0.311070793 | No |
| 6 | Endometrial | 0.227816872 | 0.033333333 | 0.341725308 | No |
| 7 | CHD | 0.432533307 | 0.038888889 | 0.556114252 | No |
| 8 | Stroke | 0.623146434 | 0.044444444 | 0.701039738 | No |
| 9 | Alzheimer | 0.951918245 | 0.05 | 0.951918245 | No |

#### Item 11.

Plot of multivariable MR results vs. univariable MR results in the UK Biobank

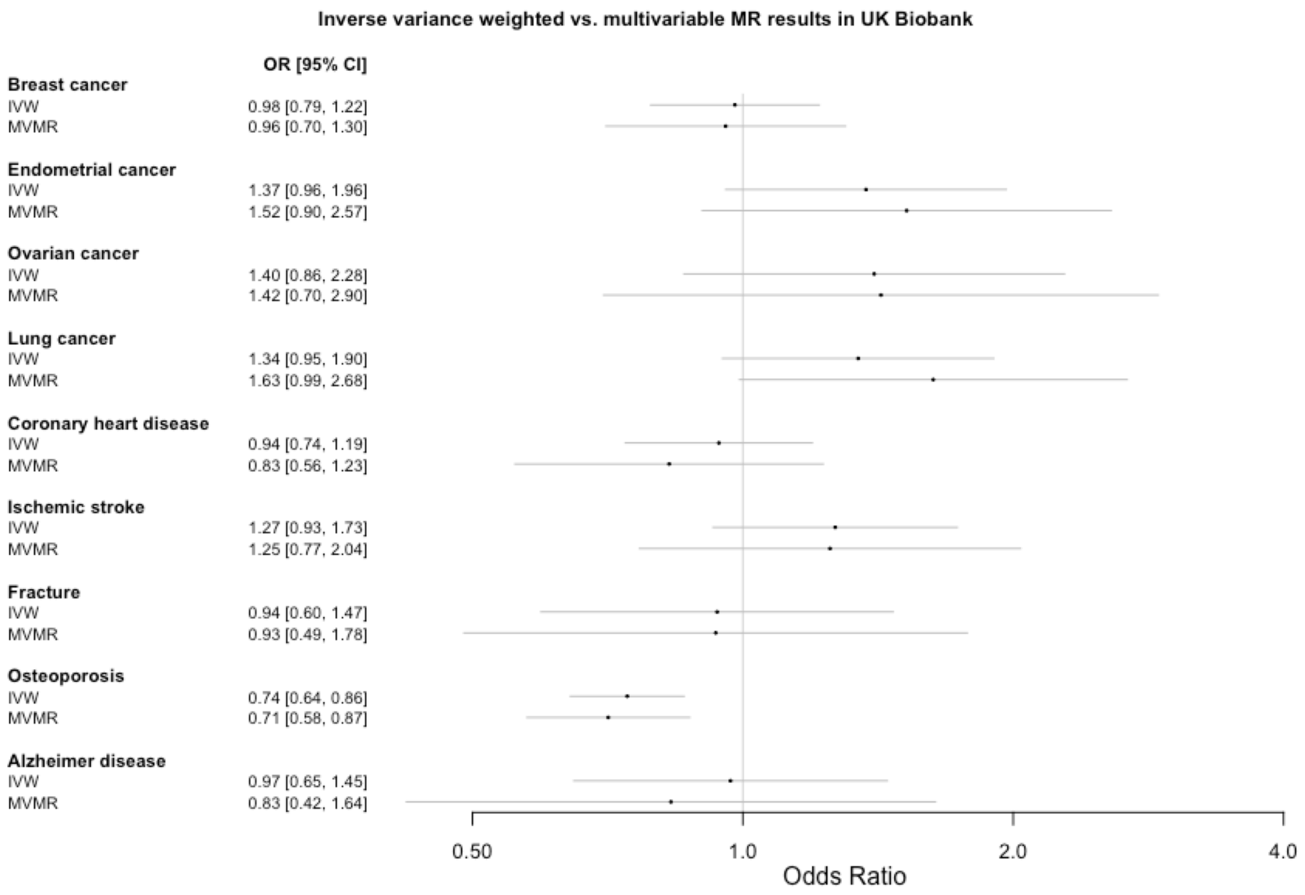

#### Item 12.

Outlier SNPs detected were rs2241584 and rs2277339. ANM = age of natural menopause; OR = odds ratio; LCL = lower confidence limit; UCL = upper confidence limit.

Supplementary Table (a) for Item 12. MR-PRESSO results in WTCCC2.

| Table. MR-PRESSO results, raw and outlier-corrected, for ischemic stroke outcome in WTCCC2. | | | | | | |
|---|---|---|---|---|---|---|
| **MR Analysis** | **P-value** | **Beta (per 5-year increase in ANM)** | **Standard error (per 5-year increase in ANM)** | **OR** | **LCL of OR** | **UCL of OR** |
| Raw | 0.98 | -0.007209415 | 0.371 | 0.99 | 0.48 | 2.05 |
| Outlier-corrected | 0.43 | 0.215126635 | 0.2693037 | 1.24 | 0.73 | 2.10 |

Supplementary Table (b) for Item 12. Meta-analysis of ischemic stroke inverse variance weighted analysis outcome substituting MR-PRESSO outlier-corrected finding from WTCCC2.

| **Data** | **OR [LCL, UCL]** |
|---|---|
| WHI | 0.58 [0.37, 0.92] |
| UKB | 1.27 [0.93, 1.73] |
| ARIC | 0.60 [0.22, 1.62] |
| CHS | 1.39 [0.60, 3.25] |
| FHS | 0.60 [0.09, 4.03] |
| MESA | 0.87 [0.09, 8.41] |
| WTCCC2 | 1.24 [0.73, 2.10] |
| **Summary for all data** | **0.96 [0.69, 1.35]** |
